# Individualized dynamic risk assessment for multiple myeloma

**DOI:** 10.1101/2024.04.01.24305024

**Authors:** Carl Murie, Serdar Turkarslan, Anoop Patel, David G. Coffey, Pamela S. Becker, Nitin S. Baliga

## Abstract

**Background:** Individualized treatment decisions for patients with multiple myeloma (MM) requires accurate risk stratification that takes into account patient-specific consequences of genetic abnormalities and tumor microenvironment on disease outcome and therapy responsiveness.

**Methods:** Previously, SYstems Genetic Network AnaLysis (SYGNAL) of multi-omics tumor profiles from 881 MM patients generated the mmSYGNAL network, which uncovered different causal and mechanistic drivers of genetic programs associated with disease progression across MM subtypes. Here, we have trained a machine learning (ML) algorithm on activities of mmSYGNAL programs within individual patient tumor samples to develop a risk classification scheme for MM that significantly outperformed cytogenetics, International Staging System, and multi-gene biomarker panels in predicting risk of PFS across four independent patient cohorts.

**Results:** We demonstrate that, unlike other tests, mmSYGNAL can accurately predict disease progression risk at primary diagnosis, pre- and post-transplant and even after multiple relapses, making it useful for individualized dynamic risk assessment throughout the disease trajectory.

**Conclusion:** mmSYGNAL provides improved individualized risk stratification that accounts for a patient’s distinct set of genetic abnormalities and can monitor risk longitudinally as each patient’s disease characteristics change.

## BACKGROUND

Multiple Myeloma (MM) is characterized by uncontrolled proliferation of malignant plasma cells originating in the bone marrow and overproduction of monoclonal immunoglobulin or M-protein. Improvements in treatments based on better knowledge of underlying pathology have improved median overall survival (OS) to >6 years.(^1^) Prognosis is primarily assessed by revised International Staging System (R-ISS)(^2^) that combines levels of β-2 microglobulin, serum albumin, lactate dehydrogenase activity, and cytogenetics at diagnosis. Fluorescence *in situ* hybridization (FISH) is used to define high-risk MM based on the presence of specific cytogenetic abnormalities, including chromosomal translocations involving the immunoglobulin heavy chain (e.g., t(4;14) and t(14;16)), and amplifications of regions involving oncogenes (amp1q) or deletions of tumor suppressors (del(17p)) that portend a shorter progression-free survival (PFS) and OS.(^3–7^) While recent advancements like Seq-Fish show promise in improving the identification of chromosomal abnormalities and merit evaluation for their potential contributions to refining risk prediction models, FISH remains the current standard in most clinical laboratories.(^8^) However, FISH testing alone is insufficient to risk stratify MM since many patients with the same cytogenetic abnormality experience varied lengths of PFS and OS, suggesting potential to further improve outcomes with finer grained risk stratification. Accordingly, many studies have used gene expression profiling to better understand subtypes and stages of MM progression.(^9–14^) In particular, two multigene biomarker panels, SKY92 (EMC-92) and GEP70 (UAMS-70), that use expression patterns of 92 and 70 genes, respectively, were commercialized into clinical tests to predict risk of disease progression.(^15, 16^) While gene expression panels are not routinely used in clinical practice they continue to play a role in advancing research efforts.(^17–19^) Recognizing their role in research, it is evident that there exists an unmet need to develop more accurate tools for risk assessment. In particular, a clinical test to longitudinally assess MM prognosis at various stages could prove transformational in improving outcomes by enabling dynamic calibration of personalized treatment plans based on the unique disease trajectory of each patient.(^20–24^)

Wall et al. advanced SYstems Genetic Network AnaLysis (SYGNAL) with Mining for Node-Edge Relationships (MINER) to analyze multi-omics data from an 881 patient cohort and construct a transcriptional regulatory network for MM (mmSYGNAL).(^25^) mmSYGNAL delineated how subsets of mutations across the patient cohort causally modulated mechanistic regulators of co-regulated genes across ∼3,000 regulons. Further, through clustering of regulons, mmSYGNAL identified 141 genetic programs whose activity profiles stratified patients into ∼25 transcriptional states that were more predictive of clinical outcome than cytogenetic subtyping. In fact, the genetic programs also uncovered subtype-specific causal and mechanistic drivers of MM progression. Additionally, mmSYGNAL also explained how patients escaped treatment and relapsed, by providing insight into mechanisms of resistance manifesting from cellular and molecular interactions within the tumor microenvironment. mmSYGNAL demonstrated, for example, that network activity of targets of FDA-approved standard of care (SOC) drugs had decreased significantly at relapse, but also suggested that MM recurrence in some patients was associated with increased sensitivity to other investigational therapies. These findings suggested that mmSYGNAL could potentially serve as a prognostic as well as a predictive tool to stratify risk and to personalize therapy regimen based on the activity profiles of genetic programs containing drug targets.

Here, we report a MM prognostic risk prediction framework based on mmSYGNAL program activity profiles within a patient’s myeloma cells. Specifically, we applied elastic net regression to identify programs within the mmSYGNAL network, whose activity profiles accurately predicted risk of disease progression in each individual across the 881 patient cohort. By training the ML algorithm on subsets of patients, we developed models that provided finer grade risk stratification within cytogenetic subtypes of MM. We have combined these models into an mmSYGNAL risk prediction framework that was tested on four independent MM cohorts (three microarray datasets from cohorts of newly diagnosed patients and one RNASeq dataset from a prospective double-blind study on a cohort of patients sampled at varied stages of the disease). These independent cohort studies demonstrated that risk prediction with mmSYGNAL significantly outperformed cytogenetics, ISS, and multi-gene biomarker panels (SKY92 and GEP70), especially across different disease stages, including primary diagnosis, pre- or post-transplant, and even after multiple relapses. Finally, we discuss how the causal and mechanistic underpinnings of genetic programs used for risk prediction also provided actionable insight into the selection of appropriate therapies for each patient.

## Materials and Methods

### Study Design

The mmSYGNAL risk prediction models were generated with a training data set and performance was analyzed with six independent validation data sets. The Interim Analysis 12 (IA12) dataset from the CoMMpass study was acquired from the Multiple Myeloma Research Foundation (MMRF) and consisted of RNASeq and cytogenetic data for 881 patients and matched clinical outcomes for 769 patients.(^26^) The mmSYGNAL model was generated with the IA12 dataset in 2021 and while the newer IA18 dataset from the same CoMMpass study did not provide sufficient numbers of new patients to justify training a new model, it did provide an independent set of relapse patients for testing model performance. The RNASeq read counts were TMM normalized, transformed into TPM values, and Z-scored by each sample and across the cohort. Seven cytogenetic abnormality risk subtypes were identified in this cohort with six based on FISH (t(4;14), t(11;14), del(17p), del(1p), del(13), amp(1q)) and one based on overexpression of *FGFR3* as a proxy for t(4;14).(^27, 28^)

Three Affymetrix data sets of 559 (GSE24080(^29^)), 282 (GSE19784(^13^)) and 426 (GSE136337(^30^)) patients with matching clinical outcomes, were normalized as described in their respective papers. A fourth dataset obtained through the Seattle Cancer Care Institute (SCCA) consisted of RNASeq, cytogenetics and clinical outcome data for 23 patients at varied disease stages.(^31^) The RNASeq data for the SCCA cohort was normalized similarly to the IA12 data set. Age and gender distributions for all data sets are shown in **Table 1**. Patients within each cohort were sub-grouped into low-, high-, and extreme-risk classes **(Table 2)** based on Guan scores **(Supplementary Methods)**.(^32^)

**Table 1:**
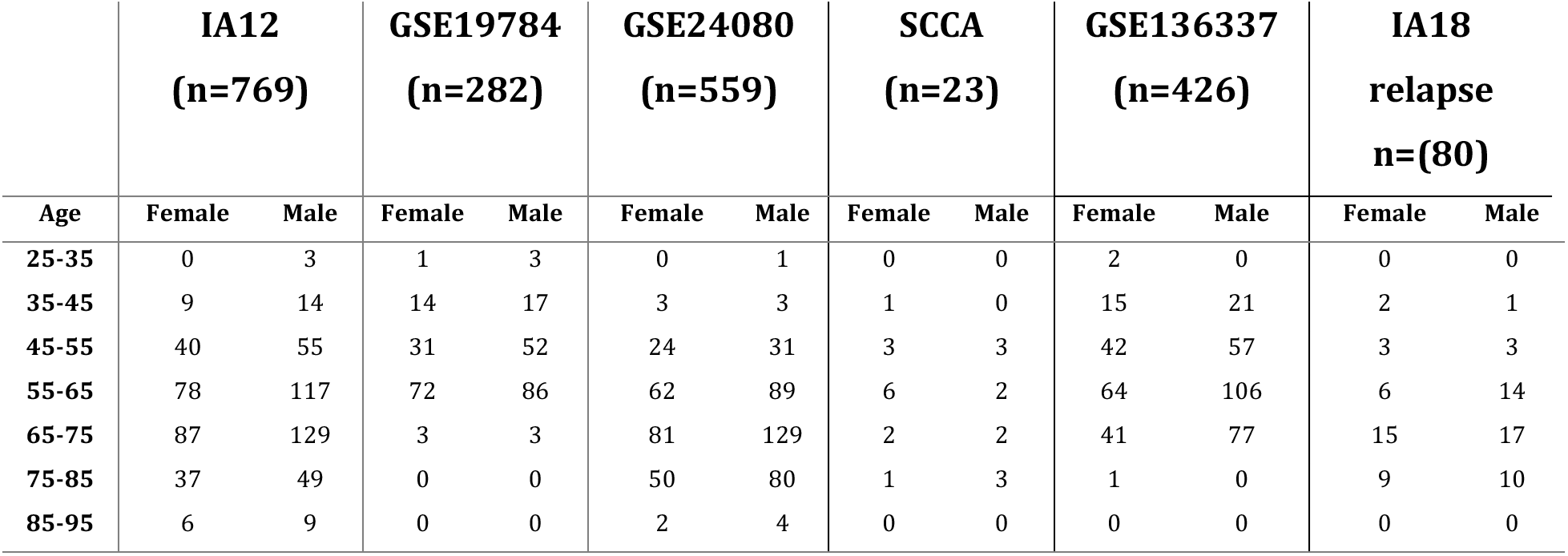
Age and gender distributions of patients in the training and validation data sets. Model was trained on the IA12 data and validated with the GSE19768, GSE24080, SCCA, GSE136337 and IA18 relapse data sets. Note that 6 patients in the IA18 relapse data set have both second- and third-line treatments and are thus repeated in the analysis which resulted in a total of 86 samples.

|  | **IA12**<br>**(n=769)** |  | **GSE19784**<br>**(n=282)** |  | **GSE24080**<br>**(n=559)** |  | **SCCA**<br>**(n=23)** |  | **GSE136337**<br>**(n=426)** |  | **IA18**<br>**relapse**<br>**n=(80)** |  |
| --- | --- | --- | --- | --- | --- | --- | --- | --- | --- | --- | --- | --- |
| **Age** | **Female** | **Male** | **Female** | **Male** | **Female** | **Male** | **Female** | **Male** | **Female** | **Male** | **Female** | **Male** |
| **25-35** | 0 | 3 | 1 | 3 | 0 | 1 | 0 | 0 | 2 | 0 | 0 | 0 |
| **35-45** | 9 | 14 | 14 | 17 | 3 | 3 | 1 | 0 | 15 | 21 | 2 | 1 |
| **45-55** | 40 | 55 | 31 | 52 | 24 | 31 | 3 | 3 | 42 | 57 | 3 | 3 |
| **55-65** | 78 | 117 | 72 | 86 | 62 | 89 | 6 | 2 | 64 | 106 | 6 | 14 |
| **65-75** | 87 | 129 | 3 | 3 | 81 | 129 | 2 | 2 | 41 | 77 | 15 | 17 |
| **75-85** | 37 | 49 | 0 | 0 | 50 | 80 | 1 | 3 | 1 | 0 | 9 | 10 |
| **85-95** | 6 | 9 | 0 | 0 | 2 | 4 | 0 | 0 | 0 | 0 | 0 | 0 |

**Table 2:**
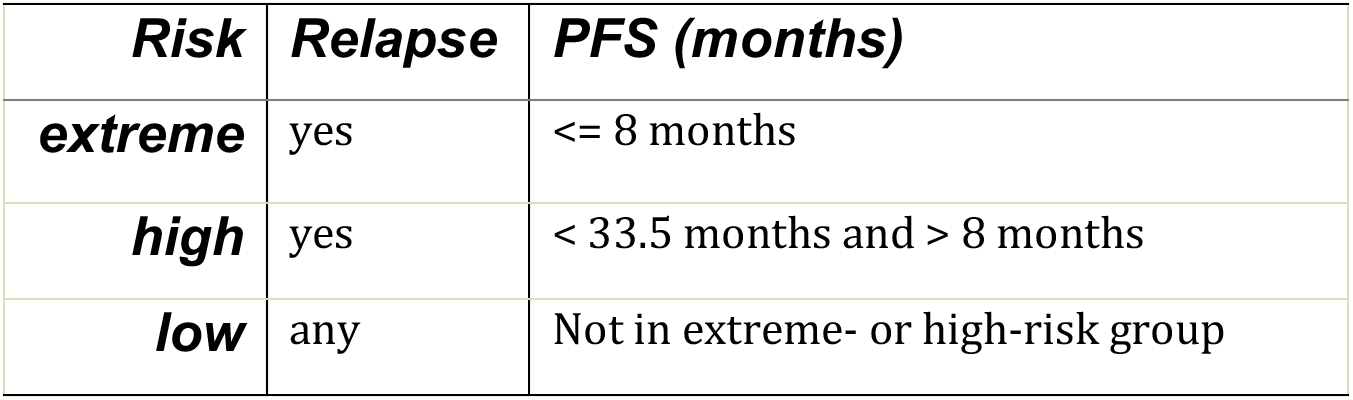
Clinical risk group PFS and relapse cutoffs generated from analysis of Guan scores. Guan scores transform survival data, PFS and relapse events, into a single numerical risk score ranging from 0 (low risk) to 1 (high risk). The inflection point for the rank ordered Guan scores for each of the three training (GSE19784, GSE24080, GSE136337) and test (IA12) data sets was identified numerically (Guan=0.5) and used as the separation point between high- and low-risk subjects. The 8 month cutoff for extreme-risk patients was based on half the median progress free survival of high-risk subjects found in Shah et al(54). The PFS values mapped from the Guan score inflection point risk cutoff were virtually identical across all data sets.

### Construction of risk prediction models based on genetic program activities within the mmSYGNAL network model

mmSYGNAL is a transcriptional regulatory network model inferred from IA12 data **(Supplementary Methods).** Gene expression and survival data from the IA12 cohort (n=769) were used to build risk prediction models for all patients (subtype-agnostic), and for the seven cytogenetic abnormality risk subtypes (t(4;14), del(1p), del(13), amp(1q), and *FGFR3* (**Table S1**). Programs were discretized as over (+1), neutral (0), or under (-1) active, based on distribution of z-scored values of member genes, as described previously.(25) Clinical outcomes were transformed to Guan score and discretized to low-, high- and extreme-risk groups (**Fig. S1**).(32) Elastic net regression (with bootstrapping and jackknife cross-validation to avoid overfitting) was applied to identify programs whose activities could stratify patients into risk classes. Training was performed on all or subsets of patients to generate sub-type agnostic or subtype-specific models, respectively, except for del(17p) and t(11;14) subtypes, which had insufficient numbers of patients for model training.(^33^) Analysis of RNASeq profiles from CD138+ myeloma cells with appropriate models was used to classify patients into low- or high-risk if the score was less (or greater) than the standard machine learning cutoff of 0.5, and as ‘extreme’ if the score was >0.6. A cutoff of 0.6 also stratified approximately ∼10% of all patients into extreme risk subgroup, which was consistent with the proportion of patients in this subgroup based on actual clinical outcome.

### Risk prediction using gene expression panels

SKY92 and GEP70 risk scores were produced with R code from the DREAM challenge(^34^), after ascertaining that our implementation reproduced Kaplan-Meier (KM) plots in the original papers (**Fig. S2**). Patients were classified as high- or low-risk if the score was higher or lower than a cutoff value reported in the original papers (SKY92=0.827 and GEP70=0.66).(^15, 16^)

### Statistical Analysis

Log-rank tests were used to evaluate the risk stratification of KM curves, and AUCs of ROC curves were used to evaluate accuracy of risk prediction. (More details on statistical analysis are in **Supplementary Information: Methods**)

## RESULTS

### mmSYGNAL scheme for predicting risk of disease progression for an individual MM patient

Previous work has shown that activity profiles of 141 programs in the mmSYGNAL network grouped 881 MM patients in the IA12 cohort into 25 transcriptional states, each associated with distinct median length of PFS.(^25^) Interestingly, patients with the same chromosomal abnormality were distributed across multiple low- and high-risk states, which indicated that cytogenetics alone was insufficient to accurately estimate risk of disease progression. Remarkably, activities of just two programs sub-stratified patients within each cytogenetic subtype (e.g., t(4;14)) into extreme (median PFS ∼5 months), high (median PFS ∼22 months) or low (median PFS ∼30 months) risk subgroups. Building on this observation, we applied elastic net regression and identified 25 programs whose activity patterns accurately stratified 769 IA12 patients into low-, high- and extreme-risk groups. This model, which was trained on all patients, will here onwards be referenced as the “subtype-agnostic model”. We investigated risk prediction on a cytogenetic subtype basis as MM is increasingly being considered as a collection of related diseases characterized by different cytogenetic abnormalities, prognoses and responses to therapy, with distinct transcriptional profiles.(^35–37^) Accordingly, for each of five cytogenetic subtypes, t(4;14), del(1p), del(13), amp(1q), and *FGFR3* (a proxy for t(4;14)), that were statistically well-represented in the patient cohort, a separate subtype-specific risk prediction model was trained (Methods). Performance of each model was evaluated on the IA12 dataset by calculating Area Under the Curve (AUC) of the Receiving Operating Characteristic (ROC) (**Fig. 1A**). Each risk model was assigned a grade based on their AUC score; t(4;14) and *FGFR3* models (AUC > 0.9) received an ‘A’ grade; amp(1q), del(1p), and del(13) (0.8 >AUC< 0.9) were assigned a ‘B’ grade, and subtype-agnostic model (AUC<0.8) was assigned a ‘C’ grade. Unsurprisingly, AUCs for combined ROC curves within A, B, and C grade models also rank ordered in a similar fashion with AUCs of 0.915, 0.848, and 0.724 respectively (**Fig. 1B**). There was significant separation between the low-, high- and extreme-risk survival curves (log rank test p-value<=1.2e-0.9) with distinct median PFS (34-44 months for the low-risk group, 19-28 months for the high-risk group and 6-14 months for the extreme-risk group; **Fig. 1C**. **Table 3**). Risk prediction using all models across the 769 patients within the IA12 dataset yielded an AUC value of 0.77 (**Fig. 1D and E)**.

**Fig 1:**
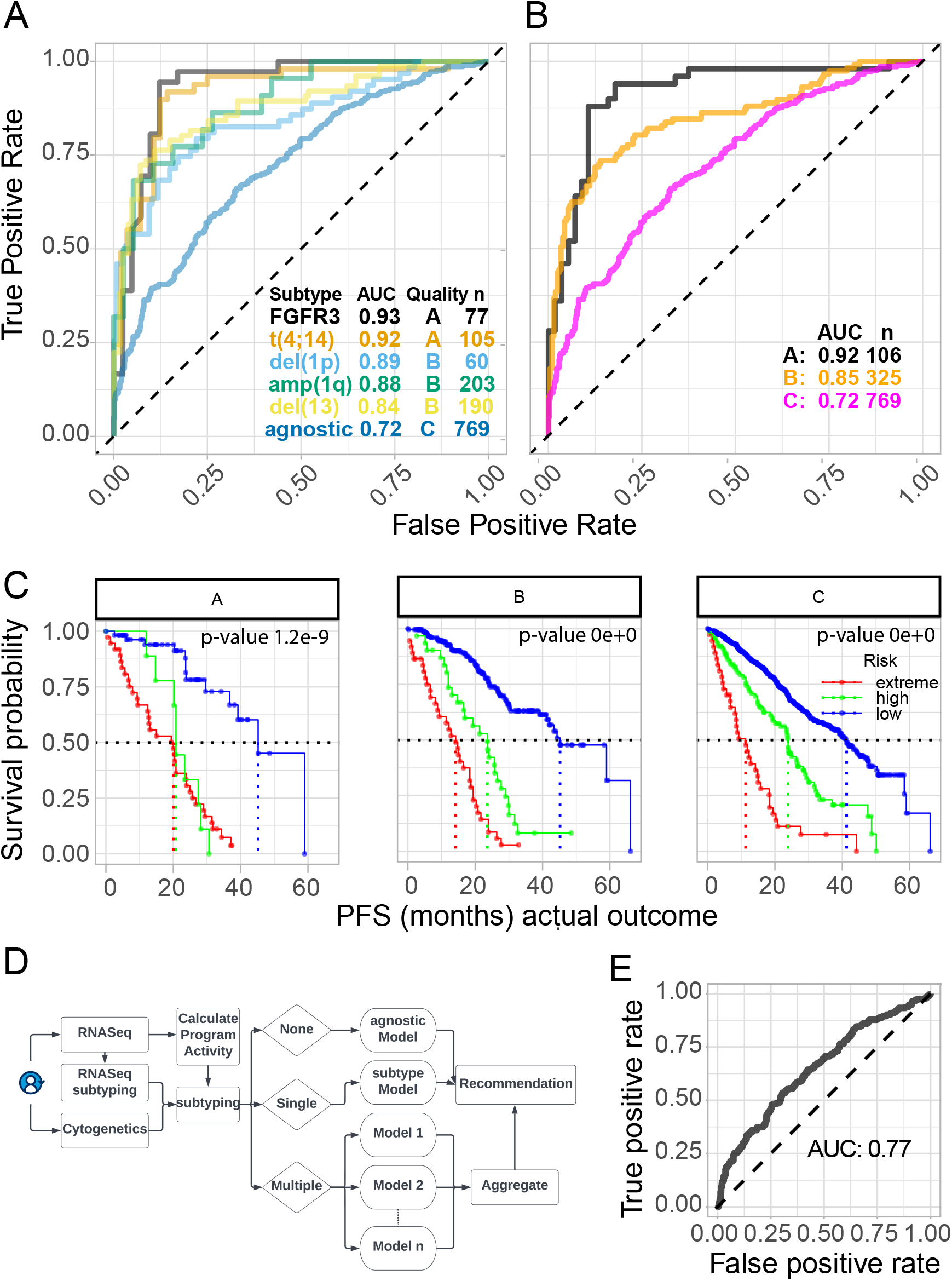
Performance of mmSYGNAL genetic subtype-specific risk and graded models. (**A**) ROC curves and sample sizes (n) of subtype specific mmSYGNAL risk models applied to 769 IA12 MM patients. Subtype risk models are organized into 3 grades based on their respective AUC scores (A >= 0.9, 0.9 < B >= 0.8, C < 0.8). (**B**) ROC curves for the IA12 patients grouped by the grade of the subtypes exhibited by each patient. A patient may be included in multiple graded groups if they exhibit multiple subtypes. If a patient exhibits 2 or more subtypes in a single grade, then the mean of the risk scores is used. (**C**) Survival plots associated with each grade with Kaplan-Meier log-rank test p-values. (**D**) Scheme for precision risk prediction for new patients using the “best” model(s). Per this scheme, the risk score of a patient that exhibits multiple subtypes is calculated using the highest-grade risk prediction model (A > B > C). For example, if a patient exhibited both the t(4;14) and the amp(1q) subtypes then the patient’s risk score would be based on t(4;14) subtype model, which had an A grade, rather than the amp(1q) subtype model, which was determined to be of B grade. If a patient exhibits multiple subtypes that are associated with equivalent graded models, then the risk score is calculated as the mean of scores generated by the highest-grade models. The patient’s risk classification defaults to the C grade subtype-agnostic model if their MM subtype is not represented by any of the subtype-specific models. (**E**) ROC curve of the mmSYGNAL best quality risk prediction scheme as described in (D) for the 769 IA12 patients. Extreme risk patients are considered as high risk in the ROC analysis.

**Table 3:** Median PFS (months) and predicted risk classification sample sizes for the three mmSYGNAL grades.

|  | *Median PFS (months)* |  |  | *n* |  |  |
| --- | --- | --- | --- | --- | --- | --- |
| *Grade* | *extreme* | *high* | *low* | *extreme* | *high* | *low* |
| **A** | 20.0 | 20.8 | 45.2 | 37 | 10 | 59 |
| **B** | 14.2 | 23.6 | 45.2 | 37 | 31 | 257 |
| **C** | 11.3 | 23.9 | 41.3 | 37 | 90 | 642 |

### mmSYGNAL significantly outperforms cytogenetics in predicting risk of disease progression in MM

Newly diagnosed MM patients with one or more high-risk chromosomal abnormalities, del(17p), t(4;14) and t(14;16), are considered by R-ISS to be at highest risk of disease progression^3^. However, there was high variability in disease outcome within cytogenetic subtypes and ISS stages across all data sets, which demonstrated that these classification methods are suboptimal in risk stratification (**Fig. S3**). We investigated the prognostic value of risk prediction based solely on cytogenetics by performing survival analysis of IA12 cohort patients stratified by the number of high-risk chromosomal abnormalities, viz. del(17p), t(4;14), t(14;16), and *FGFR3*. While the Kaplan-Meier (KM) plots showed proportional increase in risk of disease progression with the number of chromosomal abnormalities (**Fig. 2A**, **Table 4**), the number of cytogenetic abnormalities alone did a poor job of rank ordering patients on risk (AUC: 0.53) relative to the mmSYGNAL risk model (AUC: 0.65, **Fig. 2B**). While mmSYGNAL improved accuracy of predicting risk of disease progression in patients sub-grouped by numbers of high-risk cytogenetic abnormality (AUC=0.58), the overall performance was best with mmSYGNAL alone (AUC: 0.65). Finally, survival analysis also demonstrated that, relative to cytogenetic abnormalities, activities of transcriptional programs are better prognostic markers for MM (**Fig. 2C**).

**Fig. 2:**
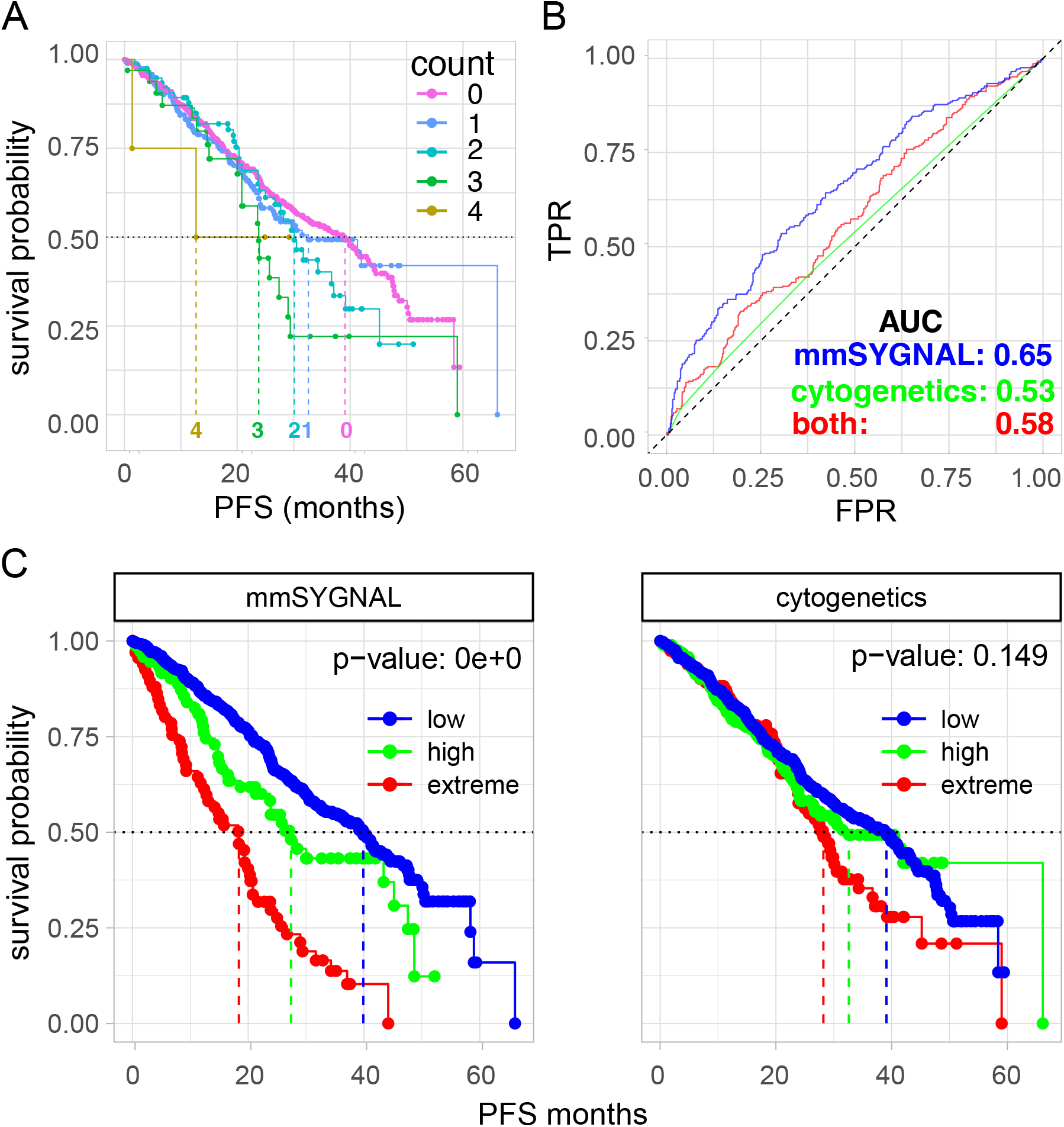
Performance of best quality mmSYGNAL model and cytogenetics. **(A**) Survival curve of risk stratification based on number of high-risk cytogenetic subtypes (t(4;14), t(4;16), del(17p), FGFR3, amp(1q)) exhibited by each patient (0 to 4). (**B**) ROC curve risk ordering based on mmSYGNAL best model, cytogenetics, and patients ordered by mmSYGNAL risk probability within each cytogenetic count group (“both”). (**C**) Survival curves and KM log-rank p-values for mmSYGNAL best model and cytogenetics risk classification (patients showing zero, one, or more than one high-risk cytogenetic abnormality are considered as low-, high- and extreme-risk groups respectively).

**Table 4:** Median PFS (months) for the high-risk cytogenetic subtype count groups.

| ***Number of abnormalities*** | ***median PFS (months)*** | ***n*** |
| --- | --- | --- |
| ***4*** | 12.7 | 5 |
| ***3*** | 23.8 | 33 |
| ***2*** | 30.1 | 81 |
| ***1*** | 32.6 | 194 |
| ***0*** | 39.1 | 456 |

### mmSYGNAL outperforms ISS and multigene biomarker panels GEP70 and SKY92

We compared performance of mmSYGNAL risk prediction to GEP70 and SKY92, which use expression levels of 70 and 92 genes, respectively, to estimate risk of disease progression.(^15, 16^) The ISS prognostic classifier (the current standard R-ISS classification was not available) was also included as it is the standard of risk assessment for newly diagnosed patients.(^2^) Accuracy of risk classification by the four approaches was tested on three cohorts (IA12, GSE19784, GSE24080). While mmSYGNAL was trained on the IA12 cohort (albeit with jackknife cross-validation to avoid overfitting), GSE19784 and GSE24080 were independent and as such served as ideal test datasets. Importantly, SKY92 was developed with GSE24080, and expected to perform best on this dataset. Moreover, both RNASeq (IA12) and microarray (GSE19784 and GSE24080) data are included allowing for assessment of model performance on disparate technology platforms.

The performance of mmSYGNAL, GEP70, and SKY92 were similar on the IA12 data set (AUCs of 0.65, 0.65, and 0.60, respectively) and GSE24080 (AUCs of 0.69, 0.70 and 0.70, respectively). Interestingly, ISS performed the worst across all data sets with AUC scores of 0.61, 0.46 and 0.64 for IA12, GSE19784 and GSE24080, respectively. While mmSYGNAL performance on GSE19784 with an AUC of 0.65 was slightly lower than GEP70 (AUC: 0.69) and SKY92 (AUC: 0.73), further analysis revealed that this might be because the two panels had identified few high-risk patients in both cohorts (**Fig. 3A**). KM survival analysis demonstrated that mmSYGNAL, SKY92 and GEP70 were all effective in classifying patients into high- and low-risk groups (**Fig. 3B)**, with median PFS values that were distinct for each dataset, but similar across all three approaches (**Table 5**). Again, ISS performance was the worst, particularly so for the GSE19784 data set with little separation between risk groups, and lower median PFS for Stage relative to Stages II and III. Importantly, only mmSYGNAL identified extreme-risk patients.

**Fig. 3:**
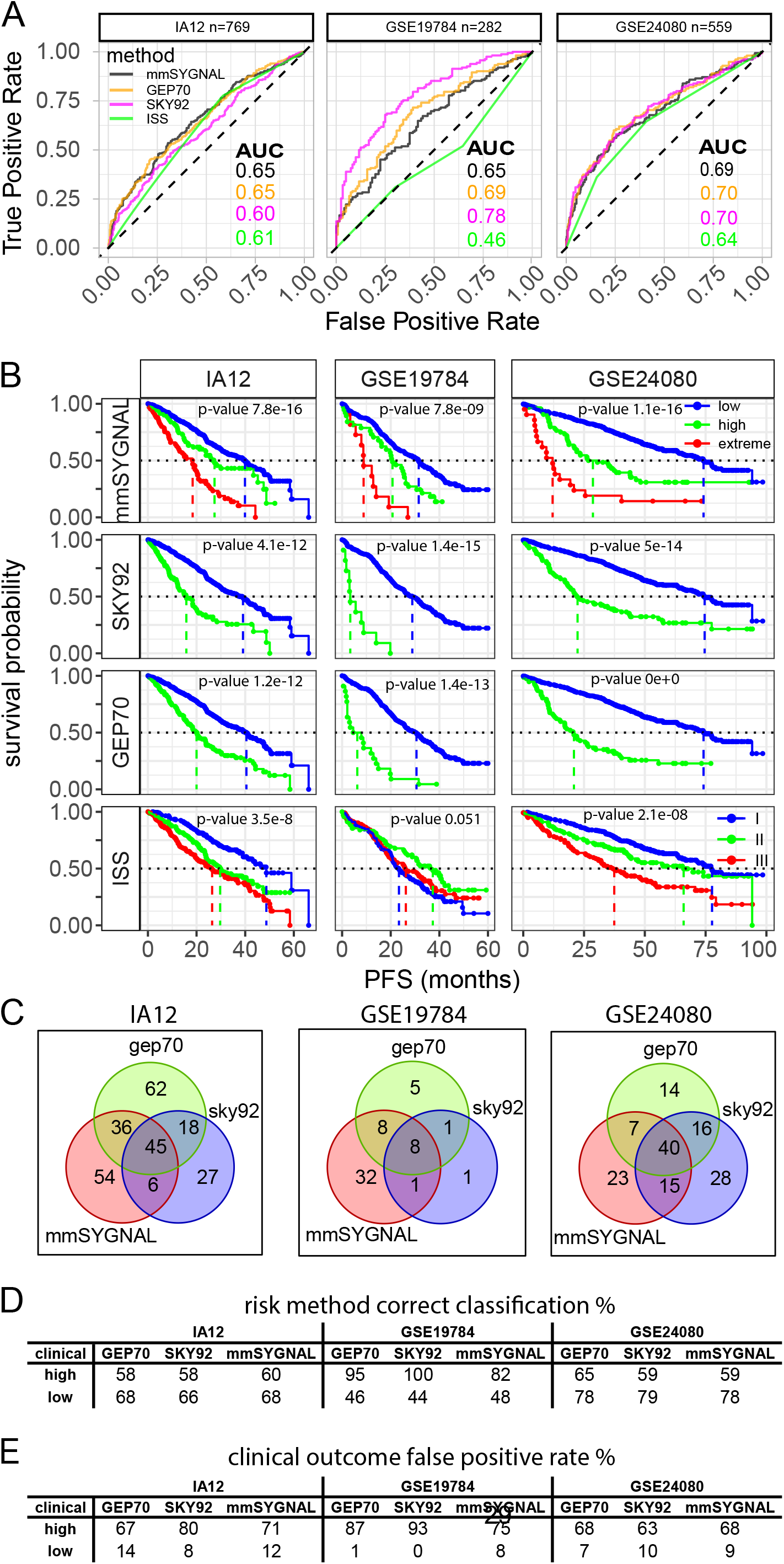
Risk prediction results for mmSYGNAL, GEP70, SKY92 and ISS. Methods were applied to the IA12 training data and two microarray validation data sets (GSE19784 and GSE24080) (**A**) ROC curves and their respective AUC scores for mmSYGNAL, GEP70 and SKY92. (**B**) Survival curves and KM log-rank p-values for mmSYGNAL, GEP70, SKY92 and ISS. All survival curves showed very low p-values. minimum p-value: 3.5e-8 except for ISS with GSE19784. (**C**) Overlap of patients that have been classified as high-risk (high- or extreme-risk for mmSYGNAL) (**D**) Percentage of correct calls made within the respective methods high- and low-risk classification groups. For example, 58% of the patients classified as high-risk by GEP70 applied to the IA12 data were also high-risk patients according to clinical outcome. (**E**) Table of false positive rates for high (high and extreme) and low risk classification groups.

**Table 5:**
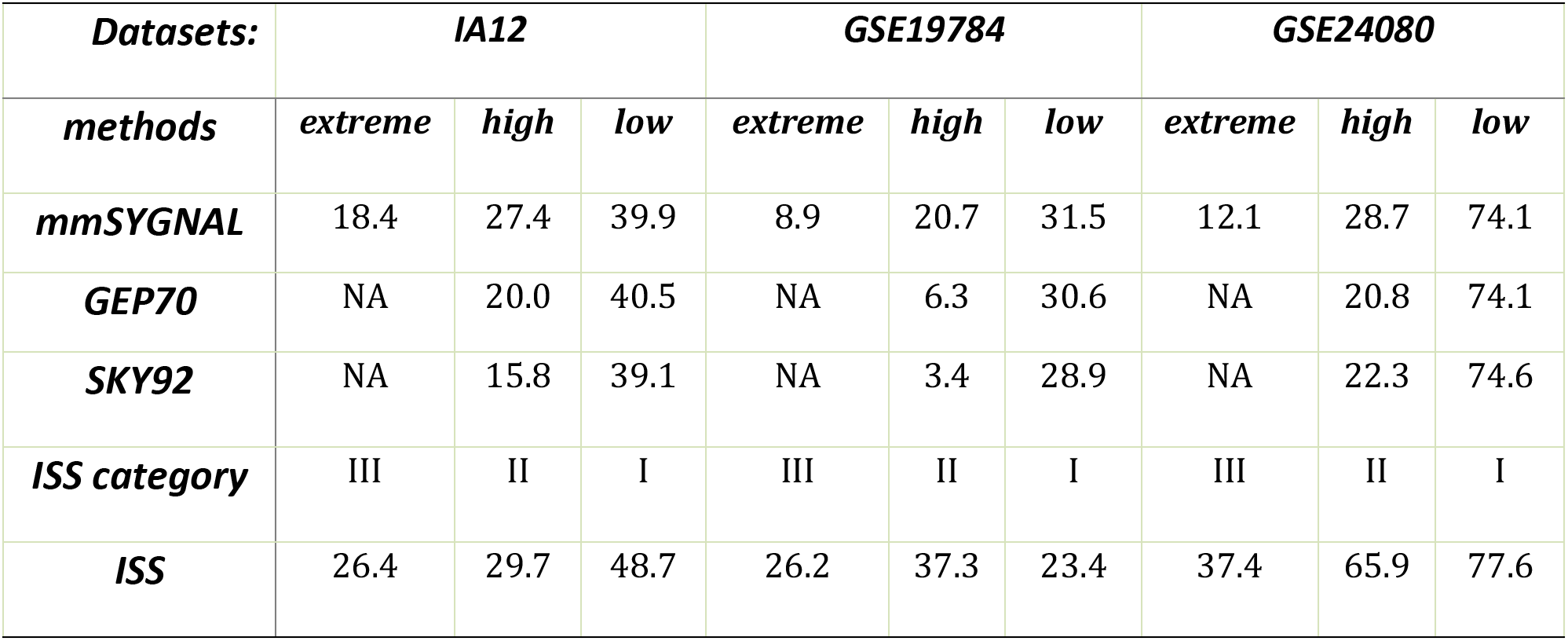
Median PFS (months) for mmSYGNAL, GEP70, and SKY92 across the IA12, GSE19784 and GSE24080 data.

| **Datasets:** | **IA12** |  |  | **GSE19784** |  |  | **GSE24080** |  |  |
| --- | --- | --- | --- | --- | --- | --- | --- | --- | --- |
| **methods** | **extreme** | **high** | **low** | **extreme** | **high** | **low** | **extreme** | **high** | **low** |
| **mmSYGNAL** | 18.4 | 27.4 | 39.9 | 8.9 | 20.7 | 31.5 | 12.1 | 28.7 | 74.1 |
| **GEP70** | NA | 20.0 | 40.5 | NA | 6.3 | 30.6 | NA | 20.8 | 74.1 |
| **SKY92** | NA | 15.8 | 39.1 | NA | 3.4 | 28.9 | NA | 22.3 | 74.6 |
| **ISS category** | III | II | I | III | II | I | III | II | I |
| **ISS** | 26.4 | 29.7 | 48.7 | 26.2 | 37.3 | 23.4 | 37.4 | 65.9 | 77.6 |

Finally, notwithstanding the significant overlap across methods, each method had identified distinct sets of high-risk patients (**Fig. 3C**). Interestingly, for the GSE19784 cohort, while SKY92 and GEP70 correctly identified only 11 and 22 high-risk patients, respectively, mmSYGNAL correctly identified 40 high-risk patients. Thus, the higher true positive rate of the two gene panels in identifying high-risk patients (95% for SKY92 and 100% for GEP70) relative to mmSYGNAL (82%) (**Fig. 3D**) came at the cost of significantly lower sensitivity, particularly so for SKY92.

### mmSYGNAL subtype-specific risk models outperform the subtype-agnostic model

Consistent with the hypothesis that MM is a collection of diseases characterized by different chromosomal abnormalities with distinct transcriptional profiles, overlapping but distinct subsets of programs contributed to risk of disease progression for each subtype of MM (**Fig. S4)**. While subtype-specific risk models significantly outperformed the subtype-agnostic model in the IA12 dataset (**Fig. 1A**), only the t(4;14) model could be evaluated on 13 and 27 t(4;14) patients in GSE19784 and GSE24080, respectively (**Table S1**). Therefore, performance of del(13) del(1p) and amp(1q) subtype-specific models were tested on a fourth dataset (GSE136337) with 77, 90, and 4 patients, respectively. While GEP70 or SKY92 risk scores could not be calculated due to unavailability of matching probe sets, the clinical metadata includes GEP70 risk classifications (high or low) and ISS stage for each patient. While all methods were effective at risk stratifying all patients (minimum p-value: 2.2e-04), the mmSYGNAL agnostic model outperformed GEP70 and ISS (**Fig. 4A-E**). Although median PFS for high- and low-risk groups (25 and 51 months respectively) were similar for the agnostic model and GEP70, only the former identified an extreme-risk (median PFS: 16 months) (**Fig. 4F**). ISS performed worst with median PFS values ranging from 30 to 58 months (Stage I to Stage III). Performance of del(13) and del(1p) subtype-specific models was even better, especially in identifying high-risk (median PFS of 21 months) and extreme-risk groups (median PFS of 9.5 and 4 months, respectively). The subtype-specific models were also better at rank ordering patients by risk, generating AUC scores of 0.62, 0.68 and 0.70 for the agnostic, del(1p) and del(13) subtypes, respectively (amp(1q) AUC was 1.0 albeit with only 4 samples). In sum, while cytogenetic abnormality alone was not a robust prognostic marker, subtype-specific mmSYGNAL models performed significantly better at predicting risk of disease progression, even relative to the subtype-agnostic risk model.

**Fig. 4:**
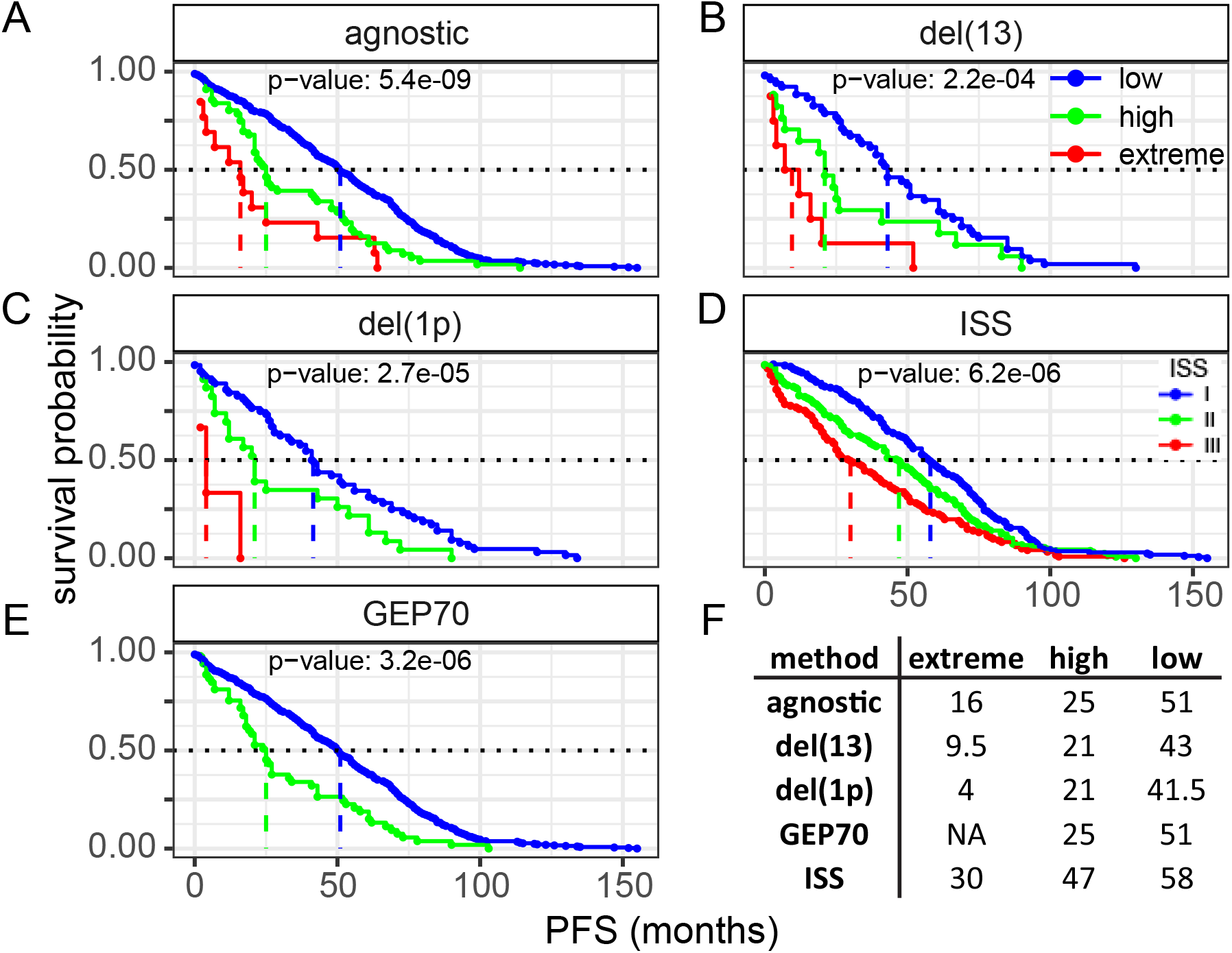
Performance of mmSYGNAL agnostic and subtype-specific models, GEP70 and ISS in the GSE136337 cohort. While we were not able to generate GEP70 or SKY92 risk scores from the processed expression data in the public repository due to unavailability of matching probe sets, the clinical metadata did include the GEP70 risk classifications (high or low) and the ISS stages for each patient and are thus shown in the analysis. Survival curves and KM log-rank p-values for (**A**) agnostic, (**B**) del(13) (**C**) ISS and (**D**) del(1p) and I GEP70 risk models. (**F**) Table of median PFS scores for all methods which correspond to the colored vertical dashed lines in the survival curves (red = extreme-risk or ISS stage III, green = high-risk or ISS stage II and blue= low-risk or ISS stage I).

### mmSYGNAL accurately predicts risk of PFS at varied disease stages, including after multiple relapses

We investigated the effectiveness of the gene panels, mmSYGNAL and ISS for longitudinal monitoring of disease progression risk beyond primary diagnosis. We applied the three risk prediction methods and ISS to 86 relapse patients in the IA18 CoMMpass dataset, who were on their second or third line of treatment. mmSYGNAL outperformed all methods in stratifying patients into low and high/extreme risk groups (KM survival curve p-values: 0.0009 (mmSYGNAL), 0.012 (GEP70), 0.076 (SKY92) and 0.020 (ISS) (**Fig. 5A**). The median length of PFS for patients classified as extreme-risk by mmSYGNAL (2.1 months) was comparable to median PFS of patients classified as high-risk by SKY92 and GEP70 (2.2 months for both). ISS stage III patients by contrast had significantly longer median PFS of 4.9 months; and there was minimal separation between survival curves for ISS Stage I (20 patients, median PFS: 8.4 months) and II (32 patient; median PFS: 8.1 months). mmSYGNAL also identified a group of high-risk patients with a median PFS of 6.1 months (**Fig. 5B**). Relative to high/extreme-risk patients, there was greater separation in mmSYGNAL survival curves for low-risk patients with median PFS of 8.1 months, as compared to median PFS of 7.4 months for SKY92 and GEP70. Importantly, mmSYGNAL identified a greater number of patients as extreme or high-risk (41 patients) than both GEP70 (18 patients), SKY92 (21 patients, **Fig. 5C**).

**Fig. 5:**
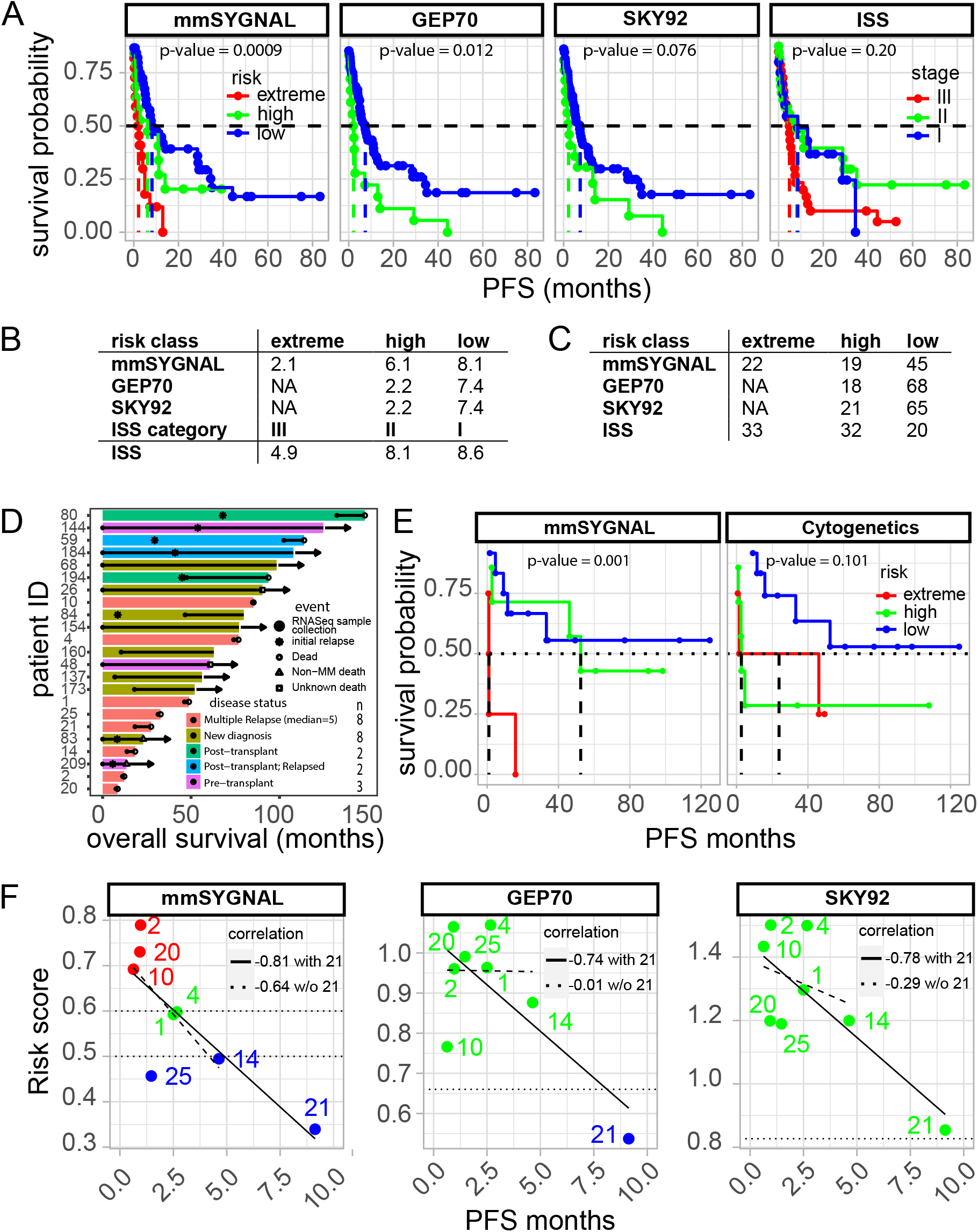
Descriptive and performance plots for mmSYGNAL, GEP70 and SKY92 applied to relapse patients. Methods were applied to 23 SCCA patients and 86 IA18 CoMMpass patients. **(A)** Survival curves for mmSYGNAL, GEP70, SKY92 and ISS applied to relapse patients from the IA18 CoMMpass trial (n=86). Tables showing the **(B)** median PFS values for each method and **(C)** sample sizes of predicted risk classifications for each method (one patient was missing ISS classification). (**D**) Swimmer plots for the SCCA patients. The arrows indicate that a patient is still going through treatment while those without arrows are either deceased or are not being followed clinically. The solid circles indicate when the RNASeq samples were obtained, and the solid lines show how long a patient was on a particular treatment before a relapse or refractory event (including death). Note that there is a mixture of samples taken at initial diagnosis and samples taken at different times along the disease trajectory. (**E**) Survival curves and KM log-rank p-values for mmSYGNAL and cytogenetics risk classification. Cytogenetic classification where low-, high- and extreme-risk categories are based on whether a patient exhibits either 0, 1, or more than one high-risk subtype, respectively. (**F**) Scatter plots and associated Pearson’s correlations of mmSYGNAL, GEP70 and SKY92 risk scores vs PFS (months) for the 8 multiple relapse/refractory patients. Patient 21’s PFS value is an outlier and thus correlations with and without that patient’s PFS identify how strongly the outlier influences correlation.

After establishing superior performance of mmSYGNAL in risk assessment of relapsed patients, we performed a double-blind study to evaluate performance of all methods across disease stages (primary diagnosis, pre- and post-transplants, and after multiple relapses). In this double-blind study, RNASeq was performed on CD138+ bone marrow mononuclear cells from 23 patients, including 8 newly diagnosed, 8 relapsed refractory (median of 5 relapses), and 7 pre- or post-transplant patients (SCCA). Notably, the cohort also represented diversity in clinical outcomes (OS: 9 -148 months and PFS: 0.7 to 52 months; **Fig. 5D**). Of all approaches tested, only mmSYGNAL was accurate in risk stratification (**Fig. 5E**). Regardless of their disease stage, mmSYGNAL stratified the 23 patients into distinct risk groups (p-value=0.001): 12 in the low-risk group, 7 in the high-risk group and 4 in the extreme-risk group. Although there was separation in survival curves of patients stratified by cytogenetics, the risk stratification was not significant (p-value=0.101), with longer median PFS for the extreme-risk group relative to the high-risk group (24 vs 3 months, respectively). By contrast, mmSYGNAL differentiated among all three risk groups with median PFS of 52 months for high-risk and 1 month for extreme-risk patients; the survival curve for the low-risk patients did not cross the 50% probability threshold (**Fig. 5E**). These results underscore the limitations of cytogenetics-based risk stratification, especially for patients with high-risk cytogenetic subtypes of MM, further motivating the need for a mmSYGNAL-type risk stratification approach.

Risk prediction by both gene expression panels performed poorly across all patients, with GEP70 calling all but one patient (P-21) high-risk, and SKY92 stratifying all patients into the high-risk group (**Fig. S5**). Interestingly, while mmSYGNAL risk scores were inversely correlated with length of PFS across all patients (r = -0.38), no correlation with PFS was observed for risk scores generated by SKY92 (r=0.1) and GEP70 (r=0.01, **Fig. S5**). Except for patient P-21, all patients with multiple relapses (P-1, P-2, P-4, P-10, P-14, P-20, P-25) were classified as high-risk by cytogenetic subtyping (**Table S2**). Yet, mmSYGNAL sub-stratified these patients into extreme-risk (P-2, P-10 and P-20), high-risk (P-1 and P-4), and low-risk (P-14, P-21 and P-25). Apart from P-25, who was misdiagnosed as low-risk, mmSYGNAL risk categorization was accurate across all three risk groups (**Fig. 5F**). ln fact, mmSYGNAL risk scores were significantly anticorrelated with length of PFS for all relapsed patients (r=-0.81), even upon excluding P-21 (r=-0.64), whose significantly longer PFS was an outlier. In contrast, the association between GEP70 and SKY92 risk scores and PFS were weaker, especially after removing P-21 (GEP70: r = -0.01; SKY92: r = -0.29). Thus, this study demonstrated the effectiveness of mmSYGNAL for longitudinally monitoring the risk of disease progression in a patient, regardless of the stage of the disease –at primary diagnosis, pre- and post-transplant, and even after multiple relapses.

### Risk-associated programs are significantly associated with targeted cancer therapies

We hypothesized that the dynamic risk assessment capability of mmSYGNAL was likely because the model was built upon causal and mechanistic principles, which could also potentially aid in drug discovery and therapy selection. To test this hypothesis, we performed survival analysis and discovered that only 25 of the total 141 programs had contributed significantly in varied weighted combinations to risk prediction by at least one model (**Fig. 6A**). While 14 programs were essential for risk prediction by a single model (e.g., Pr-98 for amp1(q) model, and Pr-110 for the subtype-agnostic model), 11 programs were important across multiple models (e.g., Pr-61 (4 models), Pr-104 (5 models) and Pr-0 (5 models)). Strikingly, under-activity of 20 programs predicted poor PFS, whereas over-activity of just 5 programs were associated with poor prognosis. The under- and over-activity of risk stratifying programs was consistent with dysregulation of similar gene sets in other cancers with poor prognosis. For example, Pr-0, a significant prognostic marker of disease progression across all subtype-specific models, was enriched for genes that have been associated with at least four other cancers (angioimmunoblastic lymphoma, leukemia, neuroblastoma, and bladder cancer).(^38–41^) Another evidence for mechanistic association of risk stratifying programs with etiology of MM was the overrepresentation of genes that are differentially regulated during early and late stages of normal differentiation from tonsil B cells -> tonsil plasma cells -> bone marrow plasma cells, with distinct expression patterns in MM plasma cells(^9^) **(Table S3, File S2**).

**Fig. 6:**
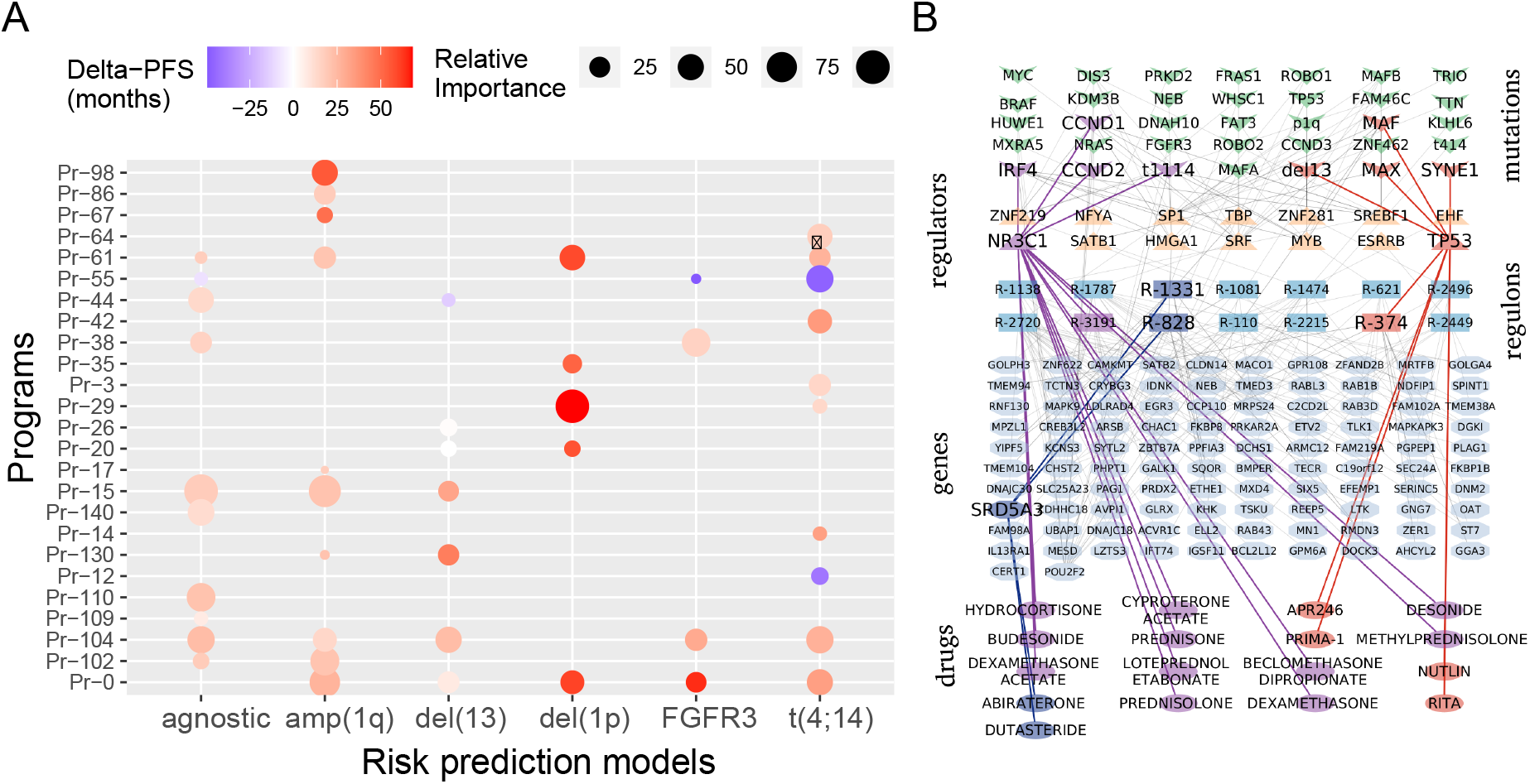
Risk-associated programs provide mechanistic insights into the biology of disease progression and therapy selection. (**A**) Relative importance of programs in risk prediction by subtype-agnostic and each subtype-specific model. Twenty-five programs that were associated with distinct survival outcomes (based on KM analysis, log-rank test p-value <0.05) contributed significantly to risk prediction by at least one model. A number of the 25 programs were significant for multiple programs which resulted in a total of 41 instances of a risk-associated program showing significance for a particular subtype. Size of each bubble is proportional to relative importance of a given program in estimating risk as determined by the scaled (0-100) absolute value of the t-statistic for each program generated by the elastic net linear regression model. Color shading of bubbles indicates the difference in median PFS of patients when the program is over-active vs. when it is under-active. For instance, Pr-29 contributed the most to the del(1p) risk model (size of bubble); median PFS of patients when this program was under-active was 50 months shorter relative to when it was over-active (dark red bubble). (**B**) The causal mechanistic flow of the regulatory network for Pr-104. The network diagram depicts SYGNAL inferred causal influences of 35 mutations on modulating 14 TFs that were implicated in mechanistic regulation of 102 genes within 14 regulons of Pr-104. Furthermore, 17 drugs in clinical trials for MM (any phase) or any other cancer (Phase IV) are shown with their respective causal flows highlighted (mutations, regulators or genes). Two causal networks show drug targets, NR3C1 (purple) and TP53 (orange), that are regulators of program Pr-104 regulons and a third where the drug target, SRD53 (yellow) is a member of two Pr-104 regulons.

If the programs were causally and mechanistically associated with disease progression, then we predicted that they should contain a significant number of targets of anticancer drugs, including MM therapies. Indeed, the 25 risk-associated programs were significantly enriched (Fischer exact test p-value=0.0003) for targets of 129 out of 399 drugs used for MM, in a Phase IV cancer trial, or in at least a Phase I MM trial. Strikingly, consistent with their mechanism of action, targets of 28 agonists and 88 antagonists were in programs associated with bad prognosis when they were under- and over-active, respectively (Fisher exact test p-value=0.0006; **Supplementary Methods)**. This list included targets of drugs used in SOC for MM, such as dexamethasone(^42, 43^) and prednisone^32,33^ both of which are agonists that target the human glucocorticoid receptor NR3C1, a TF of regulon R-3191 in Pr-104. Specifically, mmSYGNAL implicated NR3C1 in transcriptional activation of regulons within Pr-104 (Cox HR=0.66, 95% CI=(0.57, 0.77), p-value=1.510^-7^) and Pr-110 (Cox HR=0.67, 95% CI=(0.56, 0.80), p-value=9.610^-6^), and Pr-69, which was not included in the risk models but had significant risk association (Cox HR=1.52, 95% CI=(0.1.29, 0.1.81), p-value=7.410^-7^). In all three cases (Pr-104, Pr-110, Pr-69) an agonist acting on NR3C1 would differentially regulate each program in a therapeutic direction, i.e., towards a low-risk state (up-regulating Pr-104 and Pr-110 and down-regulating Pr-69) (**Fig 6B** and **Fig. S6**). Thus, these findings demonstrate the potential for leveraging the causal and mechanistic association of risk-associated programs to further develop mmSYGNAL into a predictive test for discovery of novel targets, repurposing drugs approved for other indications, and selecting therapeutic interventions based on the activity profiles of disease-driving programs in each patient.

## DISCUSSION

Personalized clinical care decisions including treatment selection for newly diagnosed MM patients is based on prognostic biomarkers such as chromosomal abnormalities that are still hindered by significant variability in survival outcomes. Given the poor accuracy of cytogenetics and the lack of use of gene expression panels there is an unmet need for improved risk assessment tools in the clinical setting. For example, in the IA12 cohort, 53% of patients with t(4;14) or *FGFR3* (high-risk chromosomal abnormalities) actually had good prognosis (long PFS). Alternatively, presumed low-risk patients with t(11;14) or no chromosomal abnormalities had significant proportions (37% and 32%, respectively) with shorter PFS.(^25^) In this regard, risk stratification based on mmSYGNAL program activities significantly improved risk stratification, even within cytogenetic subtypes, demonstrating that transcriptional states of myeloma cells are more accurate than cytogenetic abnormalities at predicting disease progression. Furthermore, our risk prediction models were tested using the IA12 data, incorporating updated cytogenetic annotations derived from Seq-FISH classifications. The results showed comparable or improved performance, underscoring the robustness and accuracy of our models (**Supplementary Information S7**). Interestingly, GEP70 and SKY92 also performed comparably across IA12, GSE19784 and GSE24080, further demonstrating that gene expression patterns do indeed contain valuable information regarding disease etiology. However, only mmSYGNAL was accurate in predicting dynamic risk across the disease trajectory of a patient.

The relatively poorer risk prediction by cytogenetics and gene expression panels, especially in later stages of MM, is likely because they are based on correlates of clinical outcomes and not directly associated with causal and mechanistic drivers of disease progression. The myriad mutations in myeloma cells of MM patients act in complex combinations that are both contextual in the bone marrow microenvironment and too large in number to model in a statistically significant manner. However, the consequences of the mutations were captured by mmSYGNAL in the architecture and activity patterns of regulons and programs that were associated with disease progression in a biologically and clinically meaningful manner.(^25^) Additionally, the improvement of cytogenetics-based risk stratification upon including program activity suggests that other mutations as well as non-genetic factors, such as the microenvironment, have major influence on disease prognosis.(^44–48^) While it is complicated to measure specific microenvironment characteristics, the activity patterns of regulons and programs within mmSYGNAL already seems to account for many of these non-genetic influences as previously demonstrated by its ability to recapitulate mechanisms of microenvironment-induced drug resistance and immune suppression within the bone marrow of relapse patients.(^25^) There is great potential to further improve the predictive power of mmSYGNAL by incorporating other risk factors considered in the R-ISS, including proportion of plasma cells and other cell types in the bone marrow aspirate and M-protein abundance –information that was unfortunately not available for any of the multiple patient cohorts analyzed in this study.

In addition to a prognostic tool for dynamic risk assessment of MM patients, there is also a need for a predictive tool that can inform clinicians on selecting non-SOC drugs matched to the characteristics of a patient’s disease. In particular, while there is broad consensus on SOC therapy for MM, there is greater uncertainty for choosing drugs for relapsed refractory MM patients.(^4, 49–51^) In this regard, mmSYGNAL could be developed further into a predictive tool for discovering and tailoring existing and new therapies to the unique biological characteristics of a patient’s disease. Our discovery that FDA-approved anti-cancer therapies (in particular SOC and investigational MM drugs) were enriched within risk-associated programs lends credibility to this strategy. Specifically, our findings suggest that activity patterns of disease-associated programs in myeloma cells could be leveraged in a rational approach to select appropriate therapies, including novel drugs. For instance, Pr-0 and Pr-86, associated with bad prognosis when under-active, were enriched in genes that were upregulated in myeloma cells exposed to Aplidin (plitidepsin, enrichment p-values: 5.37E-10 and 2.23E-2, respectively). Aplidin, a marine-derived anti-myeloma compound, inhibits proliferation of myeloma cells by inducing apoptosis(^52^) and is in Phase 3 clinical trial (NCT01102426) in combination with dexamethasone for relapsed refractory MM.(^53^) We hypothesize that high-risk patients with under-active Pr-0 and Pr-86 would likely benefit from Aplidin treatment (see **File S2** for additional examples for selecting drugs based on program activity profiles). Thus, by virtue of its causal and mechanistic association with the etiology and progression of MM, mmSYGNAL has the demonstrated utility as a prognostic tool for individualized dynamic risk assessment along the disease trajectory, and the potential for development into a predictive tool for selecting treatments that specifically target disease drivers in each patient.

## Additional Information

### Author contributions

N.S.B., P.S.B. and D.G.C conceived the project and designed experiments. P.S.B and D.G.C. provided and curated the patient-derived biopsy tumor specimens and RNA-seq data. C.M., S.T., A.P., P.S.B., D.G.C., and N.S.B. performed and/or contributed to data analysis and interpretation. C.M., S.T., A.P., P.S.B., D.G.C., and N.S.B. drafted and edited the manuscript.

### Data availability

#### Data

Microarray data are available at: GSE19784, GSE24080 and GSE136337. IA12 and IA18 data can be requested from MMRF. SCCA data can be requested from.

#### Code

mmSYGNAL risk prediction models: https://github.com/baliga-lab/mmSYGNAL-risk-prediction-models

mmSYGNAL (original code base): https://github.com/baliga-lab/miner MINER (updated interface): https://github.com/baliga-lab/miner3

#### Competing interests

NB is a co-founder and member of the Board of Directors of Sygnomics, Inc., which will commercialize the SYGNAL technology. AP and ST have equity stakes in Sygnomics, Inc. The terms of these arrangements have been reviewed and approved by ISB and Duke University in accordance with their conflict-of-interest policies. PB received institutional research support from Glycomimetics, Pfizer, Notable labs, and is an advisor to Accordant Health Services (CVS Caremark).

## Funding Information

This work was supported by a NCI-5R01AI141953-04 (N.S.B.), NSF1565166 (N.S.B.), NCI-5R01CA259469-02, NSF2042948. And Washington Research Foundation Funding (N.S.B.), a grant from the Brotman Baty Institute for Precision Medicine (UW) Catalytic Collaboration Grant (DGC and PSB), and a private donation to the University of Washington Foundation. This research was funded in part through the NIH/NCI Cancer Center Support Grant P30 CA015704 (PI Thomas Lynch, MD). The Multiple Myeloma Research Foundation generously made available the Interim Analysis 12 (IA12) version of the CoMMpass clinical trial data.

## Definitions

mmSYGNAL: multiple myeloma SYstems Genetic Network AnaLysis
MM: Multiple Myeloma
OS: Overall Survival
PFS: Progress Free Survival
FISH: Flourescent In Situ Hybridization
MINER: MIning for Node-Edge Relationships
AUC: Area Under the Curve
ROC: Reciever Operating Curve
R-ISS: Revised International Staging System
ISS: International Staging System
KM: Kaplan-Meier
IA12: Interim Analysis 12
IA18: Interim Analysis 18
GEP: Gene Expression Profile
CoMMpass: Relating <u>C</u>linical <u>O</u>utcomes in <u>MM</u> to <u>P</u>ersonal <u>Ass</u>essment of Genetic Profile
SCCA: Seattle Cancer Care Alliance
FDA: US Food and Drug Association
MMRF: Mulitple Myeloma Research Foundation
TMM: Trimmed Mean of M values
TPM: Transcript Per Million
DREAM: **D**ialogue for **R**everse **E**ngineering **A**ssessment and **M**ethods

## Supporting information

Table of GSEA cancer related results

## Data Availability

Data. Microarray data are available at: GSE19784, GSE24080 and GSE136337. IA12 and IA18 data can be requested from MMRF. SCCA data can be requested from.
Code.
mmSYGNAL risk prediction models: https://github.com/baliga-lab/mmSYGNAL-risk-prediction-models
mmSYGNAL (original code base): https://github.com/baliga-lab/miner
MINER (updated interface): https://github.com/baliga-lab/miner3

https://github.com/baliga-lab/mmSYGNAL-risk-prediction-models

https://github.com/baliga-lab/miner

https://github.com/baliga-lab/miner

## Notes

### Author Declarations

IA12 and IA18 rnaSeq data can be obtained from the MMRF (https://themmrf.org). SCCA data can be requested from.

## References

1. Rajkumar SV. Myeloma Today: Disease Definitions and Treatment Advances. American journal of hematology. 2016;91(1):90–100.

2. Rajkumar SV, Dimopoulos MA, Palumbo A, Blade J, Merlini G, Mateos M-V, et al. International Myeloma Working Group updated criteria for the diagnosis of multiple myeloma. The Lancet Oncology. 2014;15(12):e538–48.

3. Palumbo A, Avet-Loiseau H, Oliva S, Lokhorst HM, Goldschmidt H, Rosinol L, et al. Revised International Staging System for Multiple Myeloma: A Report From International Myeloma Working Group. Journal of Clinical Oncology. 2015;33(26):2863–9.

4. Kumar SK, Rajkumar V, Kyle RA, van Duin M, Sonneveld P, Mateos M-V, et al. Multiple myeloma. Nature Reviews Disease Primers. 2017;3:17046.

5. Manier S, Salem KZ, Park J, Landau DA, Getz G, Ghobrial IM. Genomic complexity of multiple myeloma and its clinical implications. Nature Reviews Clinical Oncology. 2017;14(2):100–13.

6. Sonneveld P, Avet-Loiseau H, Lonial S, Usmani S, Siegel D, Anderson KC, et al. Treatment of multiple myeloma with high-risk cytogenetics: a consensus of the International Myeloma Working Group. Blood. 2016;127(24):2955–62.

7. Hanamura I. Multiple myeloma with high-risk cytogenetics and its treatment approach. Int J Hematol. 2022;115(6):762–77.

8. Miller C YJ, Derome M, Donnely A, Marrian J, McBride K, Auclair D, Keats J. A Comparison of Clinical FISH and Sequencing Based FISH Estimates in Multiple Myeloma: An Mmrf Commpass Analysis. Blood. 2016;128(22):374.

9. Zhan F, Huang Y, Colla S, Stewart JP, Hanamura I, Gupta S, et al. The molecular classification of multiple myeloma. Blood. 2006;108(6):2020–8.

10. Agnelli L, Bicciato S, Fabris S, Baldini L, Morabito F, Intini D, et al. Integrative genomic analysis reveals distinct transcriptional and genetic features associated with chromosome 13 deletion in multiple myeloma. Haematologica. 2007;92(1):56–65.

11. Decaux O, Lodé L, Magrangeas F, Charbonnel C, Gouraud W, Jézéquel P, et al. Prediction of Survival in Multiple Myeloma Based on Gene Expression Profiles Reveals Cell Cycle and Chromosomal Instability Signatures in High-Risk Patients and Hyperdiploid Signatures in Low-Risk Patients: A Study of the Intergroupe Francophone du Myélome. Journal of Clinical Oncology. 2008;26(29):4798–805.

12. Chng WJ, Kumar S, VanWier S, Ahmann G, Price-Troska T, Henderson K, et al. Molecular Dissection of Hyperdiploid Multiple Myeloma by Gene Expression Profiling. Cancer Research. 2007;67(7):2982–9.

13. Broyl A, Hose D, Lokhorst H, de Knegt Y, Peeters J, Jauch A, et al. Gene expression profiling for molecular classification of multiple myeloma in newly diagnosed patients. Blood. 2010;116(14):2543–53.

14. Shaughnessy JD, Qu P, Usmani S, Heuck CJ, Zhang Q, Zhou Y, et al. Pharmacogenomics of bortezomib test-dosing identifies hyperexpression of proteasome genes, especially PSMD4, as novel high-risk feature in myeloma treated with Total Therapy 3. Blood. 2011;118(13):3512–24.

15. Kuiper R, Broyl A, de Knegt Y, van Vliet MH, van Beers EH, van der Holt B, et al. A gene expression signature for high-risk multiple myeloma. Leukemia. 2012;26(11):2406–13.

16. Shaughnessy JD, Zhan F, Burington BE, Huang Y, Colla S, Hanamura I, et al. A validated gene expression model of high-risk multiple myeloma is defined by deregulated expression of genes mapping to chromosome 1. Blood. 2007;109(6):2276–84.

17. Kuiper R, Zweegman S, van Duin M, van Vliet MH, van Beers EH, Dumee B, et al. Prognostic and predictive performance of R-ISS with SKY92 in older patients with multiple myeloma: the HOVON-87/NMSG-18 trial. Blood Adv. 2020;4(24):6298–309.

18. Chen YT, Valent ET, van Beers EH, Kuiper R, Oliva S, Haferlach T, et al. Prognostic gene expression analysis in a retrospective, multinational cohort of 155 multiple myeloma patients treated outside clinical trials. Int J Lab Hematol. 2022;44(1):127–34.

19. Mohan M, Weinhold N, Schinke C, Thanedrarajan S, Rasche L, Sawyer JR, et al. Daratumumab in high-risk relapsed/refractory multiple myeloma patients: adverse effect of chromosome 1q21 gain/amplification and GEP70 status on outcome. Br J Haematol. 2020;189(1):67–71.

20. van Es N. Dynamic prediction modeling for cancer-associated venous thromboembolism. Journal of Thrombosis and Haemostasis. 2020;18(6):1276–7.

21. Alexander M, Ball D, Solomon B, MacManus M, Manser R, Riedel B, et al. Dynamic Thromboembolic Risk Modelling to Target Appropriate Preventative Strategies for Patients with Non-Small Cell Lung Cancer. Cancers. 2019;11(1):50.

22. Kurtz DM, Esfahani MS, Scherer F, Soo J, Jin MC, Liu CL, et al. Dynamic Risk Profiling Using Serial Tumor Biomarkers for Personalized Outcome Prediction. Cell. 2019;178(3):699–713.e19.

23. Park Y, Kim BK, Park JY, Kim DY, Ahn SH, Han K-H, et al. Feasibility of dynamic risk assessment for patients with repeated trans-arterial chemoembolization for hepatocellular carcinoma. BMC Cancer. 2019;19:363.

24. Pitoia F, Jerkovich F. Dynamic risk assessment in patients with differentiated thyroid cancer. Endocrine-Related Cancer. 2019;26(10):R553–R66.

25. Wall MA, Turkarslan S, Wu W-J, Danziger SA, Reiss DJ, Mason MJ, et al. Genetic program activity delineates risk, relapse, and therapy responsiveness in multiple myeloma. npj Precision Oncology. 2021;5(1):1–15.

26. US National Institutes of Health. Relating clinical outcomes in multiple myeloma to personal assessment of genetic profile (CoMMpass). Clinical Trials website. https://clinicaltrials.gov/ct2/show/NCT01454297.

27. Kalff A, Spencer A. The t(4;14) translocation and FGFR3 overexpression in multiple myeloma: prognostic implications and current clinical strategies. Blood Cancer Journal. 2012;2(9):e89-e.

28. Ashby C, Boyle EM, Bauer MA, Mikulasova A, Wardell CP, Williams L, et al. Structural variants shape the genomic landscape and clinical outcome of multiple myeloma. Blood Cancer J. 2022;12(5):85.

29. Shi L, Campbell G, Jones WD, Campagne F, Wen Z, Walker SJ, et al. The MicroArray Quality Control (MAQC)-II study of common practices for the development and validation of microarray-based predictive models. Nature Biotechnology. 2010;28(8):827–38.

30. Danziger SA, McConnell M, Gockley J, Young MH, Rosenthal A, Schmitz F, et al. Bone marrow microenvironments that contribute to patient outcomes in newly diagnosed multiple myeloma: A cohort study of patients in the Total Therapy clinical trials. PLoS Med. 2020;17(11):e1003323.

31. Coffey DG, Cowan AJ, DeGraaff B, Martins TJ, Curley N, Green DJ, et al. High-Throughput Drug Screening and Multi-Omic Analysis to Guide Individualized Treatment for Multiple Myeloma. JCO precision oncology. 2021;5:PO.20.00442.

32. Huang Z, Zhang H, Boss J, Goutman SA, Mukherjee B, Dinov ID, et al. Complete hazard ranking to analyze right-censored data: An ALS survival study. PLOS Computational Biology. 2017;13(12):e1005887.

33. Zou H, Hastie T. Regularization and Variable Selection via the Elastic Net. Journal of the Royal Statistical Society Series B (Statistical Methodology). 2005;67(2):301–20.

34. bswhite. bswhite/Celgene-Multiple-Myeloma-Challenge-Baseline-Models. 2019.

35. Chng WJ, Dispenzieri A, Chim CS, Fonseca R, Goldschmidt H, Lentzsch S, et al. IMWG consensus on risk stratification in multiple myeloma. Leukemia. 2014;28(2):269–77.

36. Rajan AM, Rajkumar SV. Interpretation of cytogenetic results in multiple myeloma for clinical practice. Blood Cancer J. 2015;5(10):e365.

37. Rajkumar SV. Multiple myeloma: Every year a new standard? Hematol Oncol. 2019;37 Suppl 1(Suppl 1):62–5.

38. Piccaluga PP, Agostinelli C, Califano A, Carbone A, Fantoni L, Ferrari S, et al. Gene expression analysis of angioimmunoblastic lymphoma indicates derivation from T follicular helper cells and vascular endothelial growth factor deregulation. Cancer Research. 2007;67(22):10703–10.

39. Dürig J, Bug S, Klein-Hitpass L, Boes T, Jöns T, Martin-Subero JI, et al. Combined single nucleotide polymorphism-based genomic mapping and global gene expression profiling identifies novel chromosomal imbalances, mechanisms and candidate genes important in the pathogenesis of T-cell prolymphocytic leukemia with inv(14)(q11q32). Leukemia. 2007;21(10):2153–63.

40. Concannon CG, Koehler BF, Reimertz C, Murphy BM, Bonner C, Thurow N, et al. Apoptosis induced by proteasome inhibition in cancer cells: predominant role of the p53/PUMA pathway. Oncogene. 2007;26(12):1681–92.

41. Osman I, Bajorin DF, Sun T-T, Zhong H, Douglas D, Scattergood J, et al. Novel blood biomarkers of human urinary bladder cancer. Clinical Cancer Research: An Official Journal of the American Association for Cancer Research. 2006;12(11 Pt 1):3374–80.

42. Alexanian R, Dimopoulos MA, Delasalle K, Barlogie B. Primary dexamethasone treatment of multiple myeloma. Blood. 1992;80(4):887–90.

43. Chari A, Vogl DT, Gavriatopoulou M, Nooka AK, Yee AJ, Huff CA, et al. Oral Selinexor-Dexamethasone for Triple-Class Refractory Multiple Myeloma. N Engl J Med. 2019;381(8):727–38.

44. Neumeister P, Schulz E, Pansy K, Szmyra M, Deutsch AJ. Targeting the Microenvironment for Treating Multiple Myeloma. Int J Mol Sci. 2022;23(14).

45. Quail DF, Joyce JA. Microenvironmental regulation of tumor progression and metastasis. Nat Med. 2013;19(11):1423–37.

46. van Nieuwenhuijzen N, Spaan I, Raymakers R, Peperzak V. From MGUS to Multiple Myeloma, a Paradigm for Clonal Evolution of Premalignant Cells. Cancer Res. 2018;78(10):2449–56.

47. Pawlyn C, Morgan GJ. Evolutionary biology of high-risk multiple myeloma. Nat Rev Cancer. 2017;17(9):543–56.

48. Ghobrial IM, Detappe A, Anderson KC, Steensma DP. The bone-marrow niche in MDS and MGUS: implications for AML and MM. Nat Rev Clin Oncol. 2018;15(4):219–33.

49. Kumar S, Baizer L, Callander NS, Giralt SA, Hillengass J, Freidlin B, et al. Gaps and opportunities in the treatment of relapsed-refractory multiple myeloma: Consensus recommendations of the NCI Multiple Myeloma Steering Committee. Blood Cancer J. 2022;12(6):98.

50. Dingli D, Ailawadhi S, Bergsagel PL, Buadi FK, Dispenzieri A, Fonseca R, et al. Therapy for Relapsed Multiple Myeloma: Guidelines From the Mayo Stratification for Myeloma and Risk-Adapted Therapy. Mayo Clin Proc. 2017;92(4):578–98.

51. Hernandez-Rivas JA, Rios-Tamayo R, Encinas C, Alonso R, Lahuerta JJ. The changing landscape of relapsed and/or refractory multiple myeloma (MM): fundamentals and controversies. Biomark Res. 2022;10(1):1.

52. Delgado-Calle J, Kurihara N, Atkinson EG, Nelson J, Miyagawa K, Galmarini CM, et al. Aplidin (plitidepsin) is a novel anti-myeloma agent with potent anti-resorptive activity mediated by direct effects on osteoclasts. Oncotarget. 2019;10(28):2709–21.

53. Mitsiades CS, Ocio EM, Pandiella A, Maiso P, Gajate C, Garayoa M, et al. Aplidin, a marine organism-derived compound with potent antimyeloma activity in vitro and in vivo. Cancer Research. 2008;68(13):5216–25.

54. Shah V, Sherborne AL, Johnson DC, Ellis S, Price A, Chowdhury F, et al. Predicting ultrahigh risk multiple myeloma by molecular profiling: an analysis of newly diagnosed transplant eligible myeloma XI trial patients. Leukemia. 2020:1–6.

