## Supplementary Information for "Individualized dynamic risk assessment for multiple myeloma"

### Material and Methods

##### Supplementary information on methodology used for risk classification

The clinical outcomes data for all subjects were first transformed separately for each data set into Guan scores(1), which combine both relapse events and length of progression free survival (PFS) ref. into a linear hazard ranking that adjusts for right censorship. This transformation allows for the application of machine learning methods, such as elastic net and random forest, on survival data. The Guan scores across all subjects were rescaled to fall between 0 and 1 where 0 is lowest risk of disease progression and 1 is highest risk. **Figure S1** shows the rank ordering of the IA12 subject's Guan scores. There are two distinct distributions present with the earlier data points (blue) showing a lower slope than the later points (green and red). The inflection point separating these two sets was numerically identified and used as the cutoff point between low- and high-risk. All the subjects above the cutoff point had a relapse event and were considered high-risk while the subjects below the cutoff point were considered low-risk regardless of whether they had a relapse. An additional extreme-risk group was identified based on an approximation of Shah et al's(2) cutoff for ultra-high-risk subjects based on PFS in months. A Guan score cutoff of 0.9 was found to approximate the eight-month cutoff based on Shah et al. **Table 2** shows a summary of the metrics used for the three risk groups using the PFS values mapped from the Guan score inflection point risk cutoff.

**Supplementary data**. The Interim Analysis 12 (IA12) data set which consisted of RNASeq and cytogenetic data for 881 patients and matched clinical outcomes for 769 patients was acquired from CoMMpass study conducted by the Multiple Myeloma Research Foundation (MMRF).(3) The RNASeq data was first trimmed means of M values (TMM) normalized, transformed into transcript per million (TPM) values, and gene expression values were Z-scored by each sample and across the entire cohort. Seven cytogenetic abnormality risk subtypes were identified in this cohort with six based on FISH (t(4;14), t(11;14), del(17p), del(1p), del(13), amp(1q)) and one based on overexpression of *FGFR3* as a proxy for t(4;14).(4, 5)

Two additional Affymetrix data sets of 559 (GSE24080(6)) and 282 (GSE19784(7)) patients, including a subset of only t(4;14) and t(11;14) cytogenetic subtypes, with matching clinical outcome data, were normalized as described in their respective papers, and used to validate mmSYGNAL risk prediction models. A third independent dataset obtained through collaboration with the Seattle Cancer Care Institute (SCCA) consisted of RNASeq, cytogenetics and clinical outcome data for 23 patients at varied disease stages (“SCCA cohort”, see below).(8) The RNASeq data for the SCCA cohort was normalized similarly to the IA12 data set. A fourth Affymetrix data set (GSE136337(9)) of 426 patients, including a subset of patients exhibiting the del(13), del(1p) and amp(1q) cytogenetic subtypes, with matching clinical outcome data was also used as an independent dataset to evaluate the performance of mmSYGNAL subtype specific risk models. Only filtered gene symbol scores were provided so we could not generate GEP70 or SKY92 risk scores due to loss of information. However, the GEP70 and ISS risk classes were included in the clinical data. Age and gender distributions for the four data sets are shown in **Table 1**. Patients within each cohort were sub-grouped into low-, high-, and extreme-risk classes based on their respective Guan scores calculated separately for each data set.(1) The Guan score transforms the clinical outcome data, PFS and relapse event, into a single linear hazard ranking score. The inflection point of the rank ordered Guan scores was numerically identified and used to separate low- and high-risk patients **(Figure S1).** An additional extreme-risk group was identified based on an approximation of the cutoff from Shah et al. for ultra-high-risk subjects based on PFS (**Table 2**).(2)

**Supplementary information on construction of risk prediction models based on genetic program activities within the mmSYGNAL network model.** The mmSYGNAL model is a transcriptional regulatory network (TRN) that was generated with RNASeq and cytogenetics data for 881 patients in the IA12 dataset. The TRN was generated with a multi-step process, including (i) unsupervised clustering of RNASeq data to organize 8,549 genes into 3,203 co-regulated modules ("regulons"); (ii) mechanistic inference of 392 transcription factors (TF) implicated in the co-regulation of genes within each regulon; and (iii) causal association of 124 mutations (including chromosomal abnormalities) to regulators and their downstream target regulons.(10) Further, activity of each regulon was discretized for each patient as over (+1), neutral (0) or under (-1) active based on its ranking in the top, middle, or bottom third of the expression distribution of that regulon across all patients, as assessed by a binomial statistical test (p-value cutoff <= 0.05). Regulons with similar activity profiles across all patients were further clustered into 141 "programs", and each program was assigned an activity value in the same manner as a regulon.

Risk prediction models were generated with a regression model where activities of 141 programs for each patient were the independent (or *x)* variables and the observed clinical outcomes, classified into low- or high-risk, were the dependent (or *y*) variables. A total of 769 patients with both gene expression and survival data in the IA12 cohort were used to build risk prediction models for (i) all patients (subtype-agnostic), (ii) for each of the seven cytogenetic abnormality risk subtypes (t(4;14) [105 patients], del(1p) [60 patients], del(13) [190 patients], amp(1q) [203 patients], and *FGFR3* [77 patients],), and (iii) for subsets of patients who exhibited no subtype (subtype-none 328 patients]) (**Table S1**). Elastic net regression was applied to identify a subset of the most informative features from a large predictive feature set.(11) The elastic net model R (4.3.1) package caret (6.0-93)) was tuned with the 'train' function to optimize the elastic net parameters (α and $\lambda$)) and 1000 bootstrap iterations were applied to avoid overfitting. Additionally, jackknife or leave-one-out cross-validation was applied to the risk models as it is particularly effective in producing low biased estimates of model performance for models built with small sample sizes such as our subtype-specific models.(12) Each mmSYGNAL subtype-specific risk model consisted of a set of coefficients for the subset of programs found to be significant risk predictors. However, no risk prediction model with significant coefficients could be generated for the del(17p), t(11;14) or the 'none' risk subtypes. It is likely that there were too few samples (n=53) to generate risk prediction model for del(17p). Additionally, we hypothesize that mmSYGNAL is better able to generate risk models for subtypes associated with high-risk than for subtypes associated with low-risk groups.

We discovered that only 55 of the total 141 programs were important for risk prediction across all subtype-agnostic and subtype specific models. We further identified 25 programs that were most associated with risk of disease progression based on Kaplan Meier (KM) survival curve analysis (p-values < 0.05). Risk of disease progression for a new patient was predicted by analyzing RNASeq profile of cells from their bone marrow aspirate (ideally enriched CD138+ myeloma cells) using the appropriate cytogenetic subtype-specific or subtype-agnostic mmSYGNAL risk prediction model(s). Each model returns a value between 0 (lowest risk) and 1 (highest risk), and for patients exhibiting multiple chromosomal abnormalities, an averaged score from top performing models was used as the final risk score. A patient was classified as low- or high-risk if the score was less (or greater) than 0.5, and as 'extreme' if the score was greater than 0.6 (see Results for more details). Approximately 10% (71 out of 769) of the patients in the IA12 data set were extreme-risk patients. Accordingly, we chose 0.6 as the extreme-risk probability cutoff as this threshold predicted that approximately 10% (68 out of 769) of all patients belonged to the extreme-risk subgroup.

**Supplementary information on risk prediction using gene expression panels**. Two gene expression panels, SKY92 and GEP70, were applied to the training and validation data sets to calculate scores that estimated risk of rapid disease progression. While SKY92 uses expression patterns of 92 genes for estimating risk of disease progression, GEP70 uses expression patterns of 72 genes. Each patient was classified as high- or low-risk based on whether their score was higher or lower than a cutoff value (SKY92 = 0.827 and GEP70 = 0.66). The SKY92 and GEP70 risk scores were produced with R code implemented for the MM DREAM challenge.(13) We ascertained that our implementation of SKY92 and GEP70 applied to the GEP2658 data set reproduced KM-plots in the respective original papers (**Figure S2**).

**Supplementary information on performance evaluation of risk prediction models**.

All statistical analyses were performed in R (v4.3.1) with default parameters. Cox hazard ratios and their associated p-values were calculated with the R package "survival". Kaplan-Meier curves were also generated with the same package to evaluate risk stratification across each risk classifications by each method. The statistical significance of risk stratification of the KM curves was generated with the log-rank test where the null hypothesis is defined as no difference between the curves. ROC curves and their associated AUCs were generated (R pROC package) to compare rank ordering based on risk scores calculated by each method. Patients in each data set were first rank ordered by each method's risk score, and sensitivity and specificity values based on clinical outcome (high/low risk classifications) were then calculated for each ordered step. The sensitivity and specificity values were then plotted as an ROC curve and the AUC's for each curve were then calculated as a performance metric where an AUC of 0.5 is equivalent to random ordering and an AUC of 1 is perfect ordering. The enrichment of FDA approved therapy targets in disease relevant regulons was generated with the Fischer exact test (see Supplementary Information).

#### Supplementary information on significance on distribution of drugs mapped to risk prediction programs

Drugs were mapped to regulons and programs by first identifying the drug target(s) for each drug. Those regulons or programs that either contained or were regulated by a particular drug target are considered to be mapped to that drug.

We wanted to test whether cancer relevant drugs were enriched in the 25 risk prediction programs identified in Figure 5. A total of 399 drugs that were either standard of care or relapse/refractory multiple myeloma drugs, in a Phase IV cancer trial, or in at least a Phase I multiple myeloma had their drug targets mapped to a mmSYGNAL program. We found 129 drugs mapped to at least one of the 25 risk prediction programs while 260 mapped to a non-risk prediction program.

The Fischer exact test was applied to determine if there was a statistically significant enrichment of drugs in the mmSYGNAL risk prediction model based on the following numbers.

| | *risk prediction* | *non-risk prediction* |
|---|---|---|
| *Drugs* | 129 | 260 |
| *Programs* | 25 | 116 |

The Fischer exact test produced a p-value of 0.0003 indicating a significant enrichment of cancer and multiple myeloma drugs mapping to mmSYGNAL.

Of the drugs where we could identify direction of effect there were 28 agonist and 88 antagonist drugs mapped to the 25 risk prediction programs. The distribution of drug effect (agonist/antagonist) vs direction of network activity associated with risk of disease progression (under/over) is shown below. Note that a drug may have targets that map to multiple programs.

| | *association with risk of disease progression* | |
|---|---|---|
| *Drug effect* | *under* | *over* |
| *Agonist* | 114 | 6 |
| *antagonist* | 93 | 23 |

The Fischer exact test produced a p-value of 0.0006 indicating a significant enrichment of agonist drugs mapping to programs where risk is associated with under-activity.

### Supplementary Tables

**Table S1: Composition of the IA12, GSE19784 and GSE24080 cohort.** Composition is based on numbers and relative proportions of patients in nine cytogenetic subtypes ('agnostic' group is all patients regardless of subtype and 'none' are patients showing no cytogenetic subtypes) and three risk categories (extreme/high is a sum of extreme- and high-risk groups) based on actual clinical outcome. GSE19784 and GSE24080 only contained 4 cytogenetic subtypes.

**IA12**

| *Cytogenetic subtype* | total | extreme | high | extreme/high | low |
|---|---|---|---|---|---|
| **FGFR3** | 77 | 13% | 34% | 47% | 53% |
| **t(4;14)** | 105 | 10% | 36% | 47% | 53% |
| **agnostic** | 769 | 9% | 28% | 37% | 63% |
| **del(1p)** | 60 | 12% | 25% | 37% | 63% |
| **amp(1q)** | 203 | 9% | 28% | 37% | 63% |
| **t(11;14)** | 153 | 10% | 27% | 37% | 63% |
| **del(17p)** | 53 | 11% | 23% | 34% | 66% |
| **del(13)** | 190 | 7% | 26% | 33% | 67% |
| **none** | 328 | 8% | 25% | 32% | 68% |

**GSE19784**

| Cytogenetic subtype | total | extreme | high | extreme/high | low |
|---|---|---|---|---|---|
| **t(4;14)** | 13 | 38% | 31% | 69% | 31% |
| **agnostic** | 282 | 14% | 43% | 57% | 43% |
| **none** | 180 | 14% | 41% | 55% | 45% |
| **t(11;14)** | 61 | 10% | 38% | 48% | 52% |

**GSE24080**

| Cytogenetic subtype | total | extreme | high | extreme/high | low |
|---|---|---|---|---|---|
| **t(4;14)** | 27 | 15.% | 30.% | 44.% | 56.% |
| **agnostic** | 559 | 8.% | 20.% | 28.% | 72.% |
| **none** | 364 | 7% | 17.% | 23.% | 77.% |
| **t(11;14)** | 93 | 8.% | 15.% | 23.% | 77.% |

**Table S2: Genetic subtypes exhibited by the SCCA patients. The numbers highlighted in yellow indicate the highest-grade risk model(s) used for each patient.**

| ***Patient ID*** | ***agnostic*** | ***t(4;14)*** | ***amp(1q)*** | ***del(17p)*** | ***t(11;14)*** | ***t(14;16)*** |
|---|---|---|---|---|---|---|
| ***1*** | 0 | 0 | 1 | 0 | 0 | 0 |
| ***2*** | 0 | 1 | 0 | 0 | 0 | 0 |
| ***4*** | 0 | 0 | 1 | 0 | 1 | 0 |
| ***10*** | 0 | 1 | 1 | 1 | 0 | 0 |
| ***14*** | 0 | 0 | 1 | 0 | 0 | 0 |
| ***20*** | 0 | 0 | 1 | 0 | 1 | 0 |
| ***21*** | 1 | 0 | 0 | 0 | 0 | 0 |
| ***25*** | 0 | 0 | 1 | 1 | 1 | 0 |

**Table S3: Cancer related functional enrichments for risk-associated programs.** Functional enrichments were selected with a cutoff of an enrichment FDR score <= 0.05 (**Supplementary File S2**). The column 'Network risk association' defines the network activity associated with poor disease progression and the column 'Enrichment direction of gene expression' defines the direction of the differential expression between cancer cells in comparison to either healthy cells or cells exposed to a putative anti-cancer drug. Columns 'Program gene count' and 'Enrichment gene count' show the number of genes in the program and relevant enrichment study respectively and column 'Overlap' show the number of genes overlapping between the program and enrichment study.

| Program | Program biological functions | Network risk association | Disease | Enrichment direction of gene expression | Enrichment FDR | Overlap | Program gene count | Enrichment gene count |
|---|---|---|---|---|---|---|---|---|
| **Pr-0** | autophagy, mRNA processes, epigenetics | under-activity | angioimmunoblastic lymphoma(14) | down regulation | 1.89E-10 | 18 | 278 | 138 |
| | | | multiple myeloma(15) | down regulation | 5.37E-10 | 30 | 278 | 445 |
| | | | multiple myeloma(16) | down regulation | 4.53E-07 | 15 | 278 | 148 |
| | | | neuroblastoma(17) | down regulation | 3.26E-05 | 16 | 278 | 238 |
| | | | bladder cancer(18) | down regulation | 1.10E-02 | 16 | 278 | 427 |
| | | | leukemia(19) | down regulation | 4.30E-02 | 12 | 278 | 325 |
| **Pr-3** | | under-activity | angioimmunoblastic lymphoma(14) | down regulation | 1.42E-03 | 9 | 177 | 138 |
| **Pr-15** | | under-activity | angioimmunoblastic lymphoma(14) | down regulation | 3.71E-02 | 6 | 90 | 138 |
| **Pr-29** | lipid and adipocyte processes, cell migration | under-activity | papillary thyroid carcinoma (PTC)(20) | down regulation | 1.31E-03 | 6 | 44 | 233 |
| | | | Angioimmunoblastic lymphoma(14) | down regulation | 1.12E-03 | 6 | 44 | 138 |
| **Pr-44** | | over-activity | lung cancer(21) | up regulation | 3.89E-02 | 4 | 99 | 56 |
| **Pr-86** | cytoskeleton reorganization | under-activity | alveolar rhabdomyosarcomas (ARMS)(22) | down regulation | 4.77E-03 | 9 | 86 | 407 |
| | | | multiple myeloma(15) | down regulation | 2.23E-02 | 8 | 86 | 445 |
| **Pr-104** | | under-activity | alveolar rhabdomyosarcomas (ARMS)(22) | down regulation | 4.91E-03 | 11 | 102 | 407 |
| **Pr-110** | | under-activity | alveolar rhabdomyosarcomas (ARMS)(22) | down regulation | 2.35E-02 | 11 | 126 | 407 |

#### Supplementary Figures

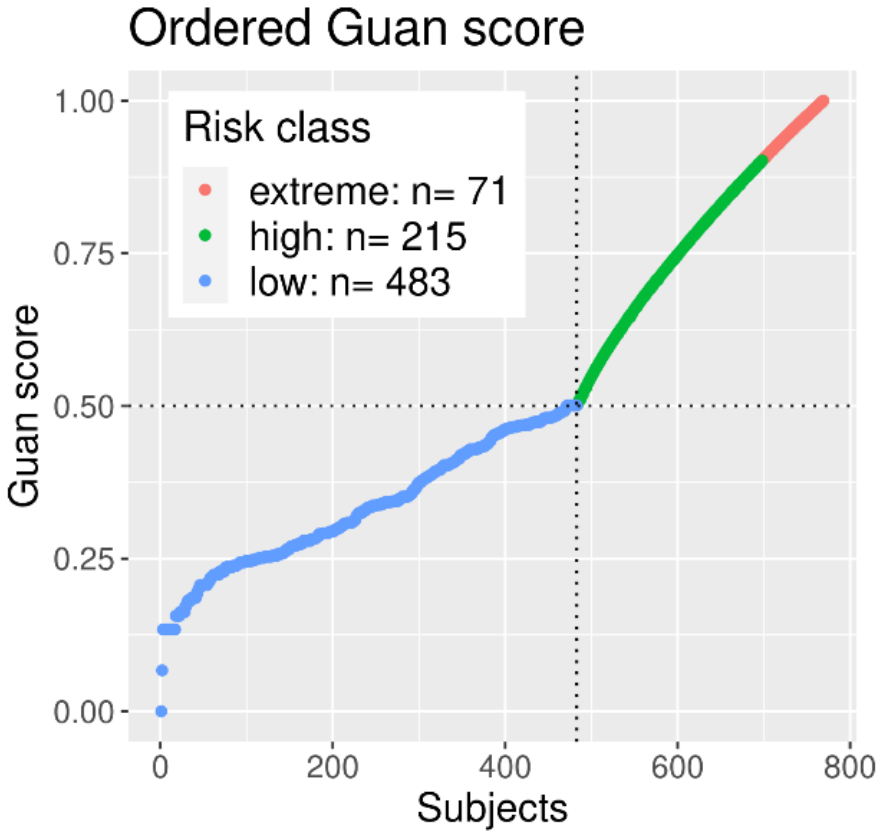


**Fig. S1: Guan scores of 769 patients ordered from low- to high-risk.** Dotted vertical and horizontal lines indicate the inflection point in guan score which was used as a threshold to separate low (blue) and high (green) risk groups. Patients that showed a relapse at less than or equal to 8 months (Guan score > 0.90) were labelled as extreme-risk (red) patients. The 8 month cutoff for extreme-risk patients was based on half the median progress free survival of high-risk subjects found in Shah et al.(2)

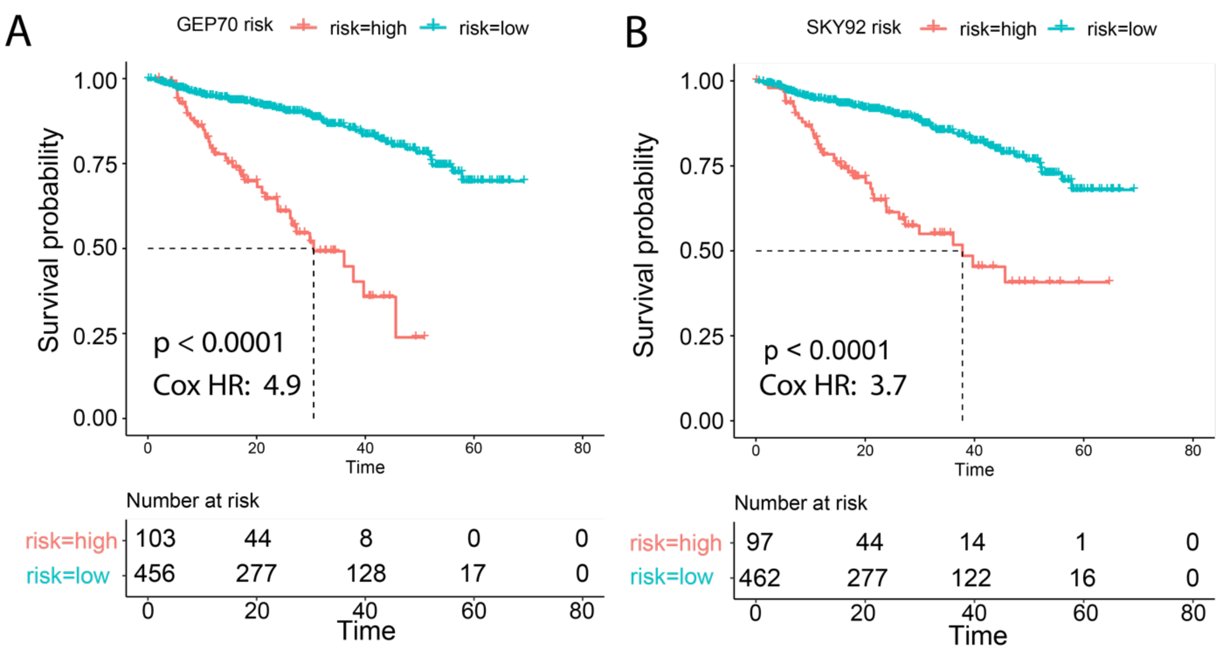


**Fig. S2.** KM curves showing performance of risk prediction by the GEP70 and SKY92 on the GSE2658 cohort. A and B. Reproduction of KM plots from the original papers, Shaugnessey et al.(23) and Kuiper et al.(24), showing the survival curves of GSE2658 patients based on risk scores generated with our implementation of the GEP70 and SKY92 gene expression panel risk score methods respectively. (A) KM plot generated from our implementation of GEP70 applied to all 559 patients in the GSE2658 cohort, since we did not have information on training and test data split. It produced a Cox HR value of 4.9 and a p-value of < 0.0001. Shaugnessey et al. showed survival curves of the training data in Figure 3C (n=351 patients, Cox HR=4.51, p-value < 0.001) and test data in Figure 4B (n=181, Cox HR=3.41, p-value < 0.001). As expected, our implementation of GEP70 reproduced results of the original paper. (B) KM plot generated from our implementation of SKY92 and its application to the entire 559 patients of the GSE2658 cohort which produced a Cox HR of 3.7 and a p-value < 0.0001. Kuiper et al. produced a KM plot of the 351 patients (training set) in Figure 1A and showed virtually identical results (Cox HR=3.4, pvalue < 0.0001) to our implementation.

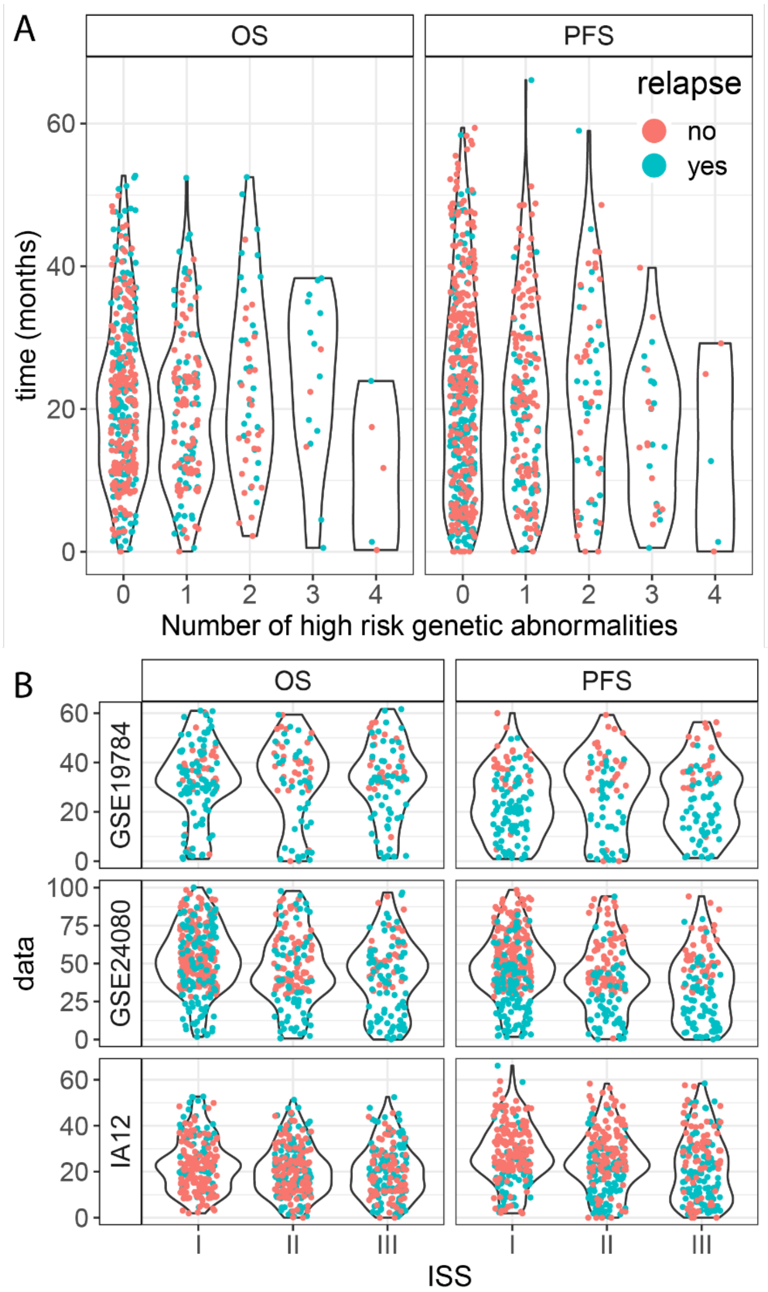


**Fig. S3: OS and PFS of validation sets.** Violin plots of OS and PFS by (**A**) number of high-risk cytogenetic abnormalities (t(4;14), del(17p), amp(1q) and FGFR3) exhibited by a patient and (**B**) ISS category. Note that there is little difference between the distributions of OS and PFS across both number of genetic abnormalities and ISS category. This shows the high variability in OS and PFS within the ISS and cytogenetic count groupings, thus demonstrating that both ISS and cytogenetics are sub-optimal in risk stratification.

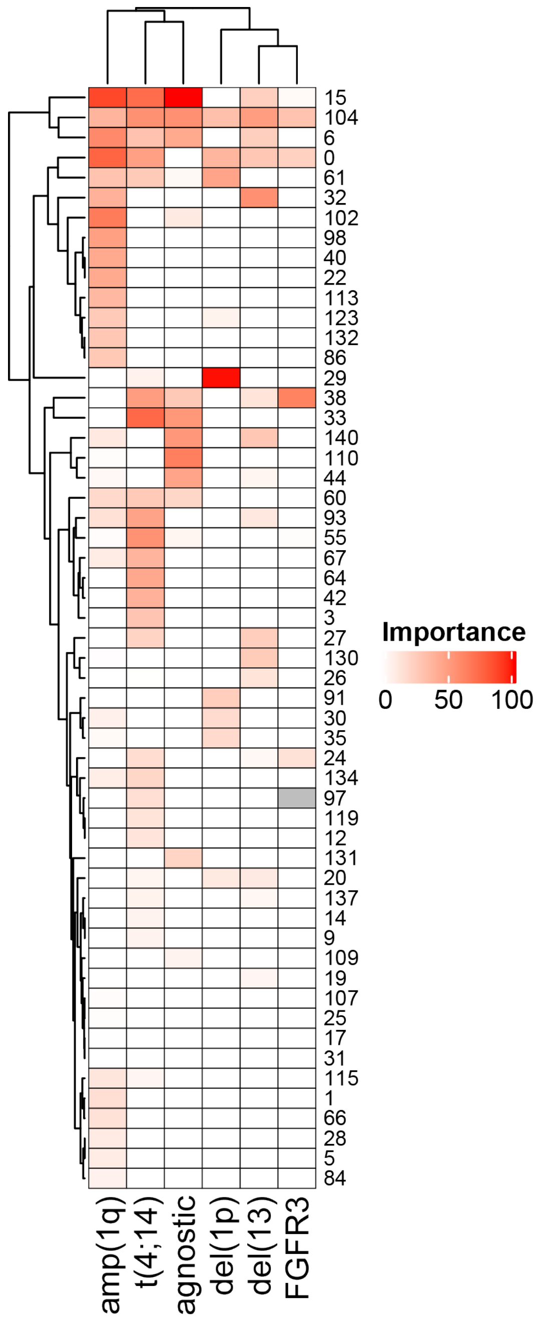


**Fig. S4: Importance scores generated by the mmSYGNAL elastic net risk prediction method.** Heat map of importance scores for all programs that were selected as significant for at least one risk prediction model. The importance scores are the scaled absolute value (0-100) of the coefficients of the elastic risk regression model. They represent the contribution of a particular program for a specific risk prediction model with 100 being the most important contributor. The white cells represent an importance of zero which indicates that the program was not used for that risk model.

Note that while there are several programs that are important across multiple subtypes, such as Pr-104, the majority are specific for a particular risk model.

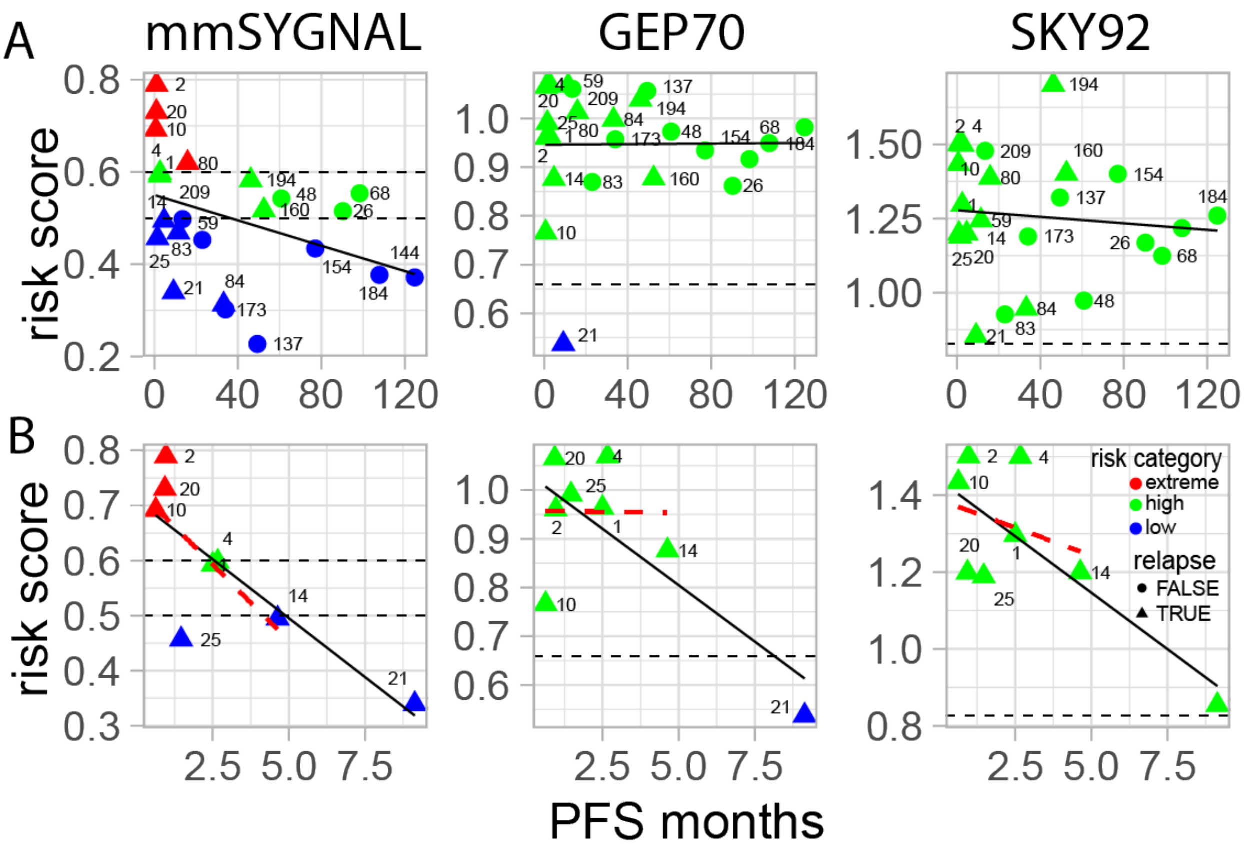


**Fig. S5: Correlation between length of PFS and risk scores generated by mmSYGNAL, GEP70 and SKY92 for 23 patients in the SCCA cohort.** Correlation plots in the top row (**A**) are for all 23 patients, and the lower row of plots (**B**) are for 8 patients with multiple relapses (median relapse = 5). Dotted horizontal lines indicate risk score thresholds used by each method for classifying low-, high- and extreme-risk groups (only mmSYGNAL identifies the extreme-risk group). Solid lines are linear regression fits to all data in each plot, while the red dashed lines are regression fits when P-21 was excluded. Triangles and circles denote patients that did or did not experience a relapse event, respectively.

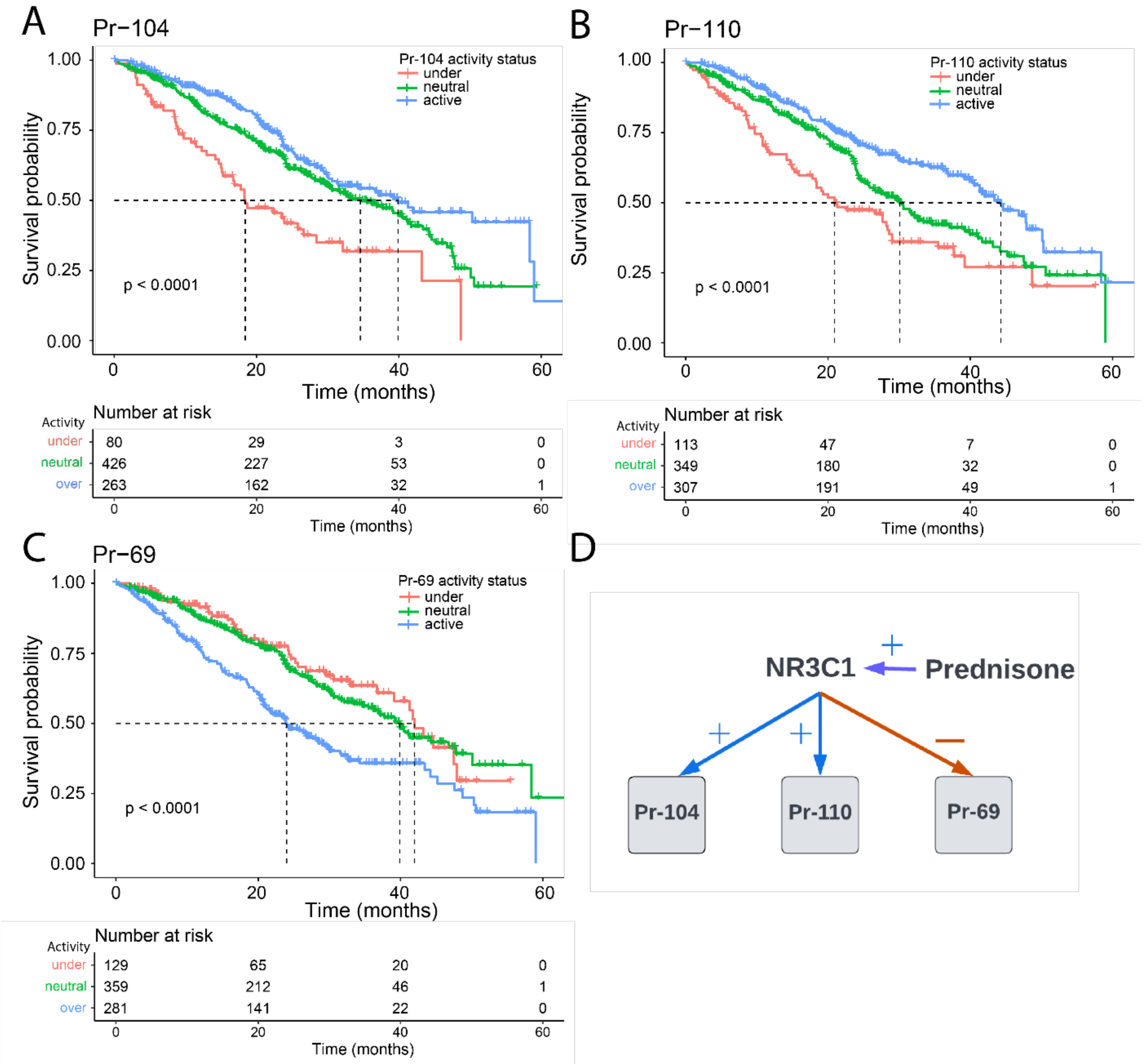


**Fig. S6: Clinical outcomes of patients stratified by activity status of three programs regulated by NR3C1.** KM survival curves for patients stratified by activity status of three programs [Pr-104 (**A**), Pr-110 (**B**) and Pr-69 (**C**)] regulated by (**D**) NR3C1, a drug target for multiple agonists including Prednisone. NR3C1 is an activator of Pr-104 and Pr-110, wherein under-activity of each program is associated with poor clinical outcomes. Triggering NR3C1 activity with an agonist will up-regulate Pr-104 and Pr-110, potentially shifting a patient to a lower risk state. Conversely, NR3C1 is a repressor of Pr-69, a program that is associated with bad prognosis when it is over-active. Activating NR3C1 will therefore repress Pr-69, rendering it neutral- or under-active and shifting a patient to a lower risk state.

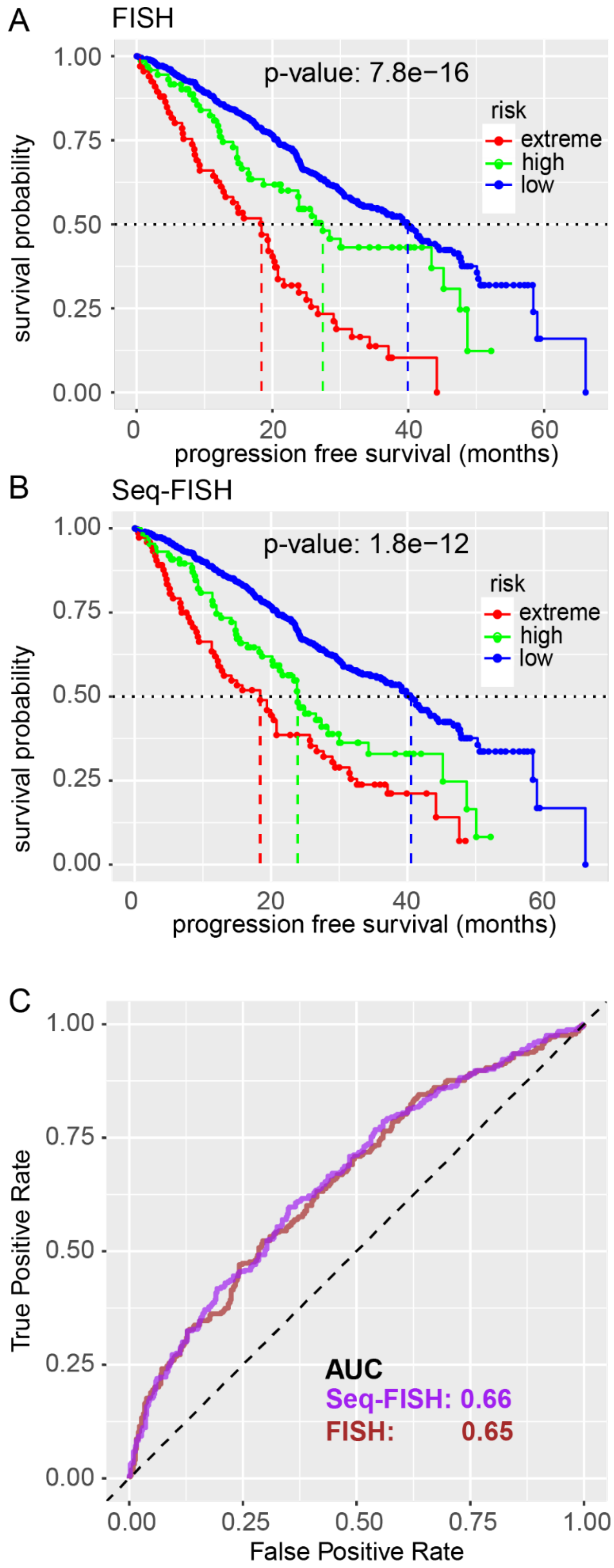
A
FISH
p-value: 7.8e−16
risk
extreme
high
low
survival probability
1.00
0.75
0.50
0.25
0.00
0
20
40
60
progression free survival (months)
B
Seq-FISH
p-value: 1.8e−12
risk
extreme
high
low
survival probability
progression free survival (months)
C
True Positive Rate
False Positive Rate
0.00
0.25
0.50
0.75
1.00
AUC
Seq-FISH: 0.66
FISH: 0.65

**Fig S7: Comparison of mmSYGNAL best quality model performance with cytogenetic subtyping based on FISH or Seq-FISH.** KM plots of the IA12 data risk prediction generated from jackknife testing with cytogenetic subtyping based on either (**A**) FISH or (**B**) Seq-FISH. (**C**) ROC curves of the IA12 risk prediction generated from jackknife testing with cytogenetic subtyping based on FISH and Seq-FISH. Seq-FISH is a whole genome sequencing data based chromosomal abnormality identifying methodology developed within MMRF.

1. Huang Z, Zhang H, Boss J, Goutman SA, Mukherjee B, Dinov ID, et al. Complete hazard ranking to analyze right-censored data: An ALS survival study. PLOS Computational Biology. 2017;13(12):e1005887.

2. Shah V, Sherborne AL, Johnson DC, Ellis S, Price A, Chowdhury F, et al. Predicting ultrahigh risk multiple myeloma by molecular profiling: an analysis of newly diagnosed transplant eligible myeloma XI trial patients. Leukemia. 2020:1-6.

3. US National Institutes of Health. Relating clinical outcomes in multiple myeloma to personal assessment of genetic profile (CoMMpass). Clinical Trials website. https://clinicaltrials.gov/ct2/show/NCT01454297.

4. Kalff A, Spencer A. The t(4;14) translocation and FGFR3 overexpression in multiple myeloma: prognostic implications and current clinical strategies. Blood Cancer Journal. 2012;2(9):e89-e.

5. Ashby C, Boyle EM, Bauer MA, Mikulasova A, Wardell CP, Williams L, et al. Structural variants shape the genomic landscape and clinical outcome of multiple myeloma. Blood Cancer J. 2022;12(5):85.

6. Shi L, Campbell G, Jones WD, Campagne F, Wen Z, Walker SJ, et al. The MicroArray Quality Control (MAQC)-II study of common practices for the development and validation of microarray-based predictive models. Nature Biotechnology. 2010;28(8):827-38.

7. Broyl A, Hose D, Lokhorst H, de Knegt Y, Peeters J, Jauch A, et al. Gene expression profiling for molecular classification of multiple myeloma in newly diagnosed patients. Blood. 2010;116(14):2543-53.

8. Coffey DG, Cowan AJ, DeGraaff B, Martins TJ, Curley N, Green DJ, et al. High-Throughput Drug Screening and Multi-Omic Analysis to Guide Individualized Treatment for Multiple Myeloma. JCO Precis Oncol. 2021;5.

9. Danziger SA, McConnell M, Gockley J, Young MH, Rosenthal A, Schmitz F, et al. Bone marrow microenvironments that contribute to patient outcomes in newly diagnosed multiple myeloma: A cohort study of patients in the Total Therapy clinical trials. PLoS Med. 2020;17(11):e1003323.

10. Plaisier CL, O'Brien S, Bernard B, Reynolds S, Simon Z, Toledo CM, et al. Causal Mechanistic Regulatory Network for Glioblastoma Deciphered Using Systems Genetics Network Analysis. Cell Systems. 2016;3(2):172-86.

11. Zou H, Hastie T. Regularization and Variable Selection via the Elastic Net. Journal of the Royal Statistical Society Series B (Statistical Methodology). 2005;67(2):301-20.

12. Friedman J, Hastie, Trevor Friedman,, Tibshirani R. The Elements of Statistical Learning. Second edition ed: Springer; 2013 November 2013.

13. bswhite. bswhite/Celgene-Multiple-Myeloma-Challenge-Baseline-Models. 2019.

14. Piccaluga PP, Agostinelli C, Califano A, Carbone A, Fantoni L, Ferrari S, et al. Gene expression analysis of angioimmunoblastic lymphoma indicates derivation from T follicular helper cells and vascular endothelial growth factor deregulation. Cancer Research. 2007;67(22):10703-10.

15. Mitsiades CS, Ocio EM, Pandiella A, Maiso P, Gajate C, Garayoa M, et al. Aplidin, a marine organism-derived compound with potent antimyeloma activity in vitro and in vivo. Cancer Research. 2008;68(13):5216-25.

16. Podar K, Raab MS, Tonon G, Sattler M, Barila D, Zhang J, et al. Up-regulation of c-Jun inhibits proliferation and induces apoptosis via caspase-triggered c-Abl cleavage in human multiple myeloma. Cancer Res. 2007;67(4):1680-8.

17. Concannon CG, Koehler BF, Reimertz C, Murphy BM, Bonner C, Thurow N, et al. Apoptosis induced by proteasome inhibition in cancer cells: predominant role of the p53/PUMA pathway. Oncogene. 2007;26(12):1681-92.

18. Osman I, Bajorin DF, Sun T-T, Zhong H, Douglas D, Scattergood J, et al. Novel blood biomarkers of human urinary bladder cancer. Clinical Cancer Research: An Official Journal of the American Association for Cancer Research. 2006;12(11 Pt 1):3374-80.

19. Dürig J, Bug S, Klein-Hitpass L, Boes T, Jöns T, Martin-Subero JI, et al. Combined single nucleotide polymorphism-based genomic mapping and global gene expression profiling identifies novel chromosomal imbalances, mechanisms and candidate genes important in the pathogenesis of T-cell prolymphocytic leukemia with inv(14)(q11q32). Leukemia. 2007;21(10):2153-63.

20. Delys L, Detours V, Franc B, Thomas G, Bogdanova T, Tronko M, et al. Gene expression and the biological phenotype of papillary thyroid carcinomas. Oncogene. 2007;26(57):7894-903.

21. Mayburd AL, Martlinez A, Sackett D, Liu H, Shih J, Tauler J, et al. Ingenuity network-assisted transcription profiling: Identification of a new pharmacologic mechanism for MK886. Clin Cancer Res. 2006;12(6):1820-7.

22. Ren Y-X, Finckenstein FG, Abdueva DA, Shahbazian V, Chung B, Weinberg KI, et al. Mouse mesenchymal stem cells expressing PAX-FKHR form alveolar rhabdomyosarcomas by cooperating with secondary mutations. Cancer Research. 2008;68(16):6587-97.

23. Shaughnessy JD, Zhan F, Burington BE, Huang Y, Colla S, Hanamura I, et al. A validated gene expression model of high-risk multiple myeloma is defined by deregulated expression of genes mapping to chromosome 1. Blood. 2007;109(6):2276-84.

24. Kuiper R, Broyl A, de Knegt Y, van Vliet MH, van Beers EH, van der Holt B, et al. A gene expression signature for high-risk multiple myeloma. Leukemia. 2012;26(11):2406-13.
